# Inpatient Antibacterial Drug Prescribing for Patients with COVID-19 in Hong Kong

**DOI:** 10.1101/2023.06.19.23291622

**Authors:** Joseph Edgar Blais, Weixin Zhang, Yun Lin, Celine SL Chui, Vincent Chi-Chung Cheng, Benjamin John Cowling, Peng Wu

## Abstract

**Background:** Hong Kong experienced four epidemic waves caused by the ancestral strain of SARS-CoV-2 in 2020-2021 and a large Omicron wave in 2022. Few studies have assessed antibacterial drug prescribing for COVID-19 inpatients throughout the pandemic.

**Objectives:** To describe inpatient antibacterial drug prescribing for COVID-19 patients throughout the pandemic and to determine factors associated with their prescription.

**Methods:** This cohort study used electronic health records of COVID-19 cases admitted to public hospitals in Hong Kong from 21 January 2020 to 30 September 2022. We assessed the prevalence and rates of inpatient antibacterial drug use, using days of therapy/1000 patient days (DOT/1000PD), and examined the association of baseline factors and disease severity with receipt of an inpatient antibacterial drug prescription.

**Results:** Among 65,810 inpatients, 54.0% were prescribed antibacterial drugs at a rate of 550.5 DOT/1000PD. Antibacterial use was lowest during wave 4 (28.0%; 246.9 DOT/1000PD), peaked in early wave 5 (64.6%; 661.2 DOT/1000PD), and then modestly declined in late wave 5 (43.2%; 464.1 DOT/1000PD) starting on 23 May 2022.

Older age, increased disease severity, and residing in an elderly care home were strongly associated with increased odds of prescription, while receiving ≥ 2 doses of COVID-19 vaccines and pre-admission use of coronavirus antivirals were associated with lower odds.

**Conclusions:** The rate of inpatient antibacterial prescribing initially declined during the pandemic, but increased during the Omicron wave when hospital capacity was overwhelmed. Despite the availability of COVID-19 vaccines and antiviral drugs, antibacterial drug use among COVID-19 inpatients remained high into late 2022.

**HIGHLIGHTS:**

- The prevalence of antibacterial drug use in hospitalized COVID-19 cases in Hong Kong declined gradually during the first four COVID-19 epidemic waves to 28.0%, but increased to 64.6% with the spread of the Omicron variant in early 2022.
- The majority of antibacterial drug prescriptions were for Access and Watch drugs, with limited use of combination therapy or macrolides.
- Older age and more severe disease were strongly associated with an inpatient antibacterial drug prescription, while vaccination and initiation of COVID-19-specific antivirals reduced the odds of antibacterial prescription.
- Despite moderate-to-high levels of vaccine coverage and the availability of antiviral drugs, 43% of COVID-19 inpatients still received antibacterial drugs in late 2022.

## INTRODUCTION

Early in the coronavirus disease 2019 (COVID-19) pandemic, concern for bacterial co-infections led to widespread prescribing of antibacterial drugs for patients with COVID-19 admitted to hospital. The overall prevalence of antibacterial prescribing was 75%, and it was 100% for patients admitted to intensive care units (ICU).[1–3] However, only 8% of hospitalized patients with COVID-19 had a confirmed bacterial co-infection at admission, with up to 20% being diagnosed with a secondary bacterial infection later during admission.[4–6] While a declining temporal trend in antibacterial drug prescribing among patients with COVID-19 early in the pandemic, few studies have investigated antibacterial drug use after widespread vaccination and the availability of oral antiviral drugs for individuals at high-risk of severe clinical outcomes, particularly during epidemics caused by Omicron variants, which emerged in late 2021.[1,3]

Antibacterial drug prescribing among inpatients with COVID-19 could contribute to the development and spread of antimicrobial resistance (AMR) to a wider population.[7] In Hong Kong, recent reports have suggested an increased number of inpatients with methicillin-resistant *Staphylococcus aureus* bacteremia, an outbreak of *Candida auris*, and a rising number of patients carrying carbapenem-resistant Enterobacteriaceae discharged from hospital to residential care homes for the elderly, indicating a worsening AMR situation during the COVID-19 pandemic.[8,9]

In Hong Kong, stringent pandemic containment measures throughout 2020 and 2021 suppressed local transmission of severe acute respiratory syndrome coronavirus 2 (SARS-CoV-2).[10–12] During this period, all persons with laboratory-confirmed infections, including asymptomatic cases, were strictly isolated in public hospitals.[13,14] The largest epidemic wave in Hong Kong was wave 5, caused by Omicron subvariants that emerged in late 2021, resulting in over one million confirmed cases and nearly 10,000 deaths within 2 months.[15] During the peak of wave 5, public hospitals were overloaded with COVID-19 cases, most of which involved older adults.

The World Health Organization (WHO) declared end of COVID-19 as a public health emergency in early May 2023. Quantifying antibacterial drug prescribing practices for COVID-19 patients throughout the pandemic allows us to establish a baseline benchmark for ongoing monitoring of antibacterial drug use and to identify potential gaps in evidence to practice. Therefore, this study aimed to describe the inpatient prescribing of antibacterial drugs over time and to assess factors associated with inpatient antibacterial use among patients with COVID-19 admitted to all public hospitals in Hong Kong up to 30 September 2022.

## METHODS

### Data sources and ethical approval

Since the start of the COVID-19 pandemic, all officially reported COVID-19 cases have been tracked by the Centre for Health Protection (CHP) of the Hong Kong Department of Health. Data from the CHP, including age, sex, confirmation date, case classification (locally acquired or imported), and COVID-19 vaccination records were linked using a pseudonymous identifier to electronic health records obtained from the Hospital Authority (HA), the statutory body responsible for providing public health care in the territory. Data obtained from the HA included patient demographics, prescription dispensing records (outpatient and inpatient), COVID-19 condition status, inpatient transaction records (dates of admission, discharge, and transfer and ward location), inpatient diagnoses, laboratory tests, and outpatient clinic attendance dates and diagnoses. In this study, we analyzed data sets from CHP and HA extracted on 30 November 2022.

The study was approved by the institutional review board of The University of Hong Kong/ Hospital Authority Hong Kong West Cluster. Data used for this analysis was part of the public health response to the pandemic, and informed consent was waived from individual patients.

### Population and setting

This cohort study included all patients with community-acquired COVID-19 who were admitted to public hospitals in Hong Kong. Community-acquired COVID-19 was defined as patients with COVID-19 who had a confirmation date of SARS-CoV-2 infection that occurred between 7 days prior to the admission date and 3 days after the admission date (**Supplementary Figure 1**). To permit sufficient follow-up time after hospital admission, patients with a COVID-19 confirmation date between 21 January 2020 and 30 September 2022 were eligible for inclusion. Patients without an inpatient admission, inpatients not discharged by 30 October 2022, patients with a missing date of birth, and imported COVID-19 cases were excluded. For patients with multiple COVID-19 related inpatient admissions episodes, we restricted our analyses to the first episode closest to their confirmation date.

### Baseline variables and disease severity definitions

Baseline variables included demographics, medical conditions, drugs, COVID-19 vaccination, and records for a microbiological culture result (see **Supplementary Table 1** for details). Dates of each epidemic wave were defined based on the inpatient admission date and categorized as follows: waves 1-2 (1 January 2020 to 30 June 2020), wave 3 (1 July 2020 to 31 October 2020), wave 4 (1 November 2020 to 30 December 2021), early wave 5 (5E, 31 December 2021 to 22 May 2022) and late wave 5 (5L, 23 May 2022 to 30 September 2022).[16] Patients were classified into one of four disease severity groups: fatal was defined as inpatient death; critical was defined as any day present in intensive care unit/high dependency unit (hereafter ICU), or requiring intubation, mechanical ventilation, or extracorporeal membrane oxygenation, or in shock; severe was defined as a prescription for remdesivir, baricitinib, tocilizumab, or dexamethasone, or requiring oxygen supplementation of 3 liters per minute or more, or an arterial blood gas saturation of 90% or less; and mild to moderate was defined as not being classified into fatal, critical, or severe disease severity (**Supplementary Table 2**).

### Antibacterial drug use and outcomes

We quantified all inpatient prescriptions for drugs listed in the British National Formulary (BNF) section 5.1: Antibacterial drugs. Antibacterial drugs were further grouped according to the WHO AWaRe classification (2021) into access, watch, reserve, and not recommended groups.[17] Among patients prescribed an antibacterial drug, we extracted the initiation day of antibacterial drug(s) for each patient and classified the earliest inpatient prescription of antibacterial drug into either monotherapy (defined as the prescription of only one antibacterial drug on the initiation day of antibacterial treatment) or combination therapy (defined as the prescription of two or more antibacterial drugs on the initiation day).

Measures of drug use followed published recommendations for in hospital antibacterial drug utilization research.[18] The prevalence of antibacterial drug prescription in all patients with community-acquire COVID-19 during follow-up was measured as the proportion of patients with any dispensed antibacterial drug prescription from the admission date to the end of follow-up. We examined the number of days of antimicrobial therapy (DOT) for each antibacterial drug and calculated the proportion of total DOT for each antibacterial drug and AWaRe group. DOT per 1000 patient days (PD) and DOT per 1000 patients were used as the main quantity metrics of antibacterial drug use.

To minimize bias caused by outliers with extremely long hospital stays, we censored the calculation of the time period at risk at 90 days. The follow-up period for an inpatient prescription ended therefore on the earliest of the discharge date, date of death, or inpatient day 89. Clinical outcomes reported include the number of days present in hospital and death during follow-up (i.e., fatal severity).

### Statistical analysis

Rates of antibacterial drug use were calculated by summing the number of DOT during a specified time period divided by the denominator (either patient days or patients). We used a logistic regression model to assess the associations between baseline variables and disease severity and the outcome of an inpatient antibacterial drug prescription, by estimating conditional odds ratios (ORs) and 95% confidence intervals. Analyses were conducted using R version 4.2.0 (R Foundation for Statistical Computing, Vienna, Austria).

## RESULTS

As of 30 September 2022, there were 1,765,405 officially confirmed COVID-19 cases in Hong Kong. Our analyses focused on 65,810 patients with COVID-19 who were likely infected in the local community (excluding imported cases and nosocomial cases) and admitted into a public hospital for isolation or treatment. All the included patients had either been discharged from or died in hospital at the time of analysis (**Supplementary Figure 2**). Over half of the patients (37,991, 57.7%) were admitted during wave 5E, which was dominated by the Omicron BA.2 subvariant (**Figure 1A**). The median age of the patient cohort was 70.0 years (interquartile range [IQR]: 44.0-84.0), with 7759 (11.8%) patients aged < 20 years (**Table 1**). The majority of patients had mild to moderate infections, with 1691 (2.6%) patients admitted to ICU. In total, 7473 (11.4%) patients died in the hospital, with 90.0% (n=6699) of inpatient deaths occurred during wave 5E.

**Figure 1.**
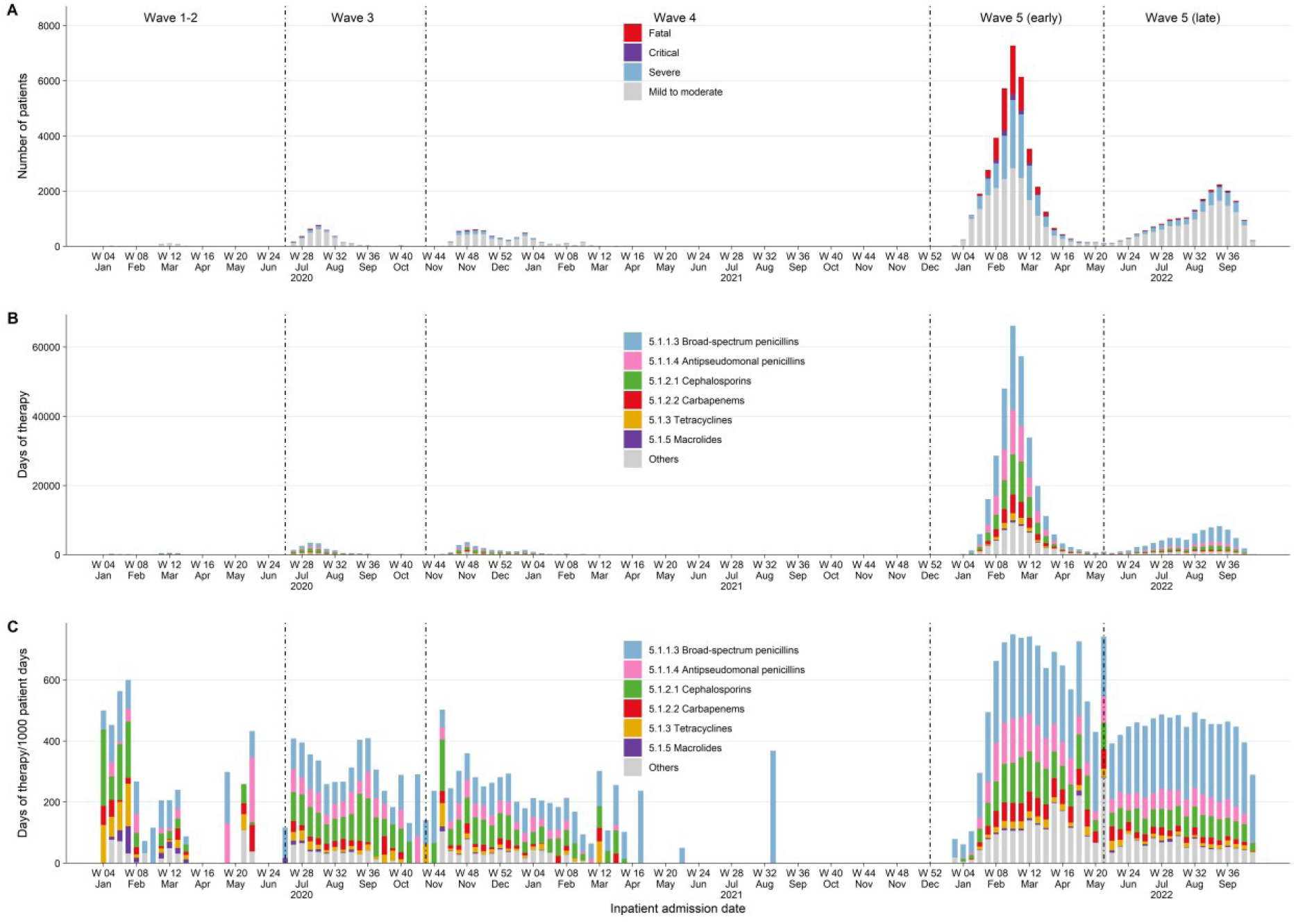
**A**, weekly number of inpatients with COVID-19 included in the study. Weekly antibacterial drug use according to BNF section in (**B**) days of therapy and (**C**) days of therapy/1000 patient days.

**Table 1.** Baseline characteristics and disease severity of patients hospitalized with confirmed COVID-19 in all public hospitals stratified by epidemic wave

| Characteristic | Wave 1-2 | Wave 3 | Wave 4 | Wave 5E | Wave 5L | Overall |
| --- | --- | --- | --- | --- | --- | --- |
| Patients | 405 (0.6) | 3402 (5.2) | 5448 (8.3) | 37991 (57.7) | 18564 (28.2) | 65810 (100.0) |
| **Demographics** |  |  |  |  |  |  |
| Sex = F/M | 191/214 (47.2/52.8) | 1765/1637 (51.9/48.1) | 2861/2587 (52.5/47.5) | 18110/19881 (47.7/52.3) | 8967/9597 (48.3/51.7) | 31894/33916 (48.5/51.5) |
| Age, years, median [IQR] | 39.0 [31.0, 55.0] | 50.0 [33.0, 62.0] | 47.0 [32.0, 61.0] | 75.0 [57.0, 87.0] | 70.0 [35.0, 81.0] | 70.0 [44.0, 84.0] |
| Age group, years |  |  |  |  |  |  |
| < 20 | 12 (3.0) | 281 (8.3) | 508 (9.3) | 3228 (8.5) | 3730 (20.1) | 7759 (11.8) |
| 20-59 | 324 (80.0) | 2042 (60.0) | 3407 (62.5) | 7124 (18.8) | 3243 (17.5) | 16140 (24.5) |
| 60-79 | 62 (15.3) | 892 (26.2) | 1328 (24.4) | 11762 (31.0) | 6435 (34.7) | 20479 (31.1) |
| ≥ 80 | 7 (1.7) | 187 (5.5) | 205 (3.8) | 15877 (41.8) | 5156 (27.8) | 21432 (32.6) |
| Old age home resident | 0 (0.0) | 0 (0.0) | 0 (0.0) | 11986 (31.5) | 1274 (6.9) | 13260 (20.1) |
| **Inpatient admission characteristics** |  |  |  |  |  |  |
| Patient days, median [IQR] | 23.0 [16.0, 30.0] | 12.0 [8.0, 17.0] | 13.0 [9.0, 17.0] | 8.0 [4.0, 14.0] | 7.0 [3.0, 11.0] | 8.0 [4.0, 14.0] |
| ICU admission | 31 (7.7) | 163 (4.8) | 232 (4.3) | 1004 (2.6) | 261 (1.4) | 1691 (2.6) |
| Patient days, median [IQR]<sup>a</sup> | 28.0 [20.0, 45.0] | 27.0 [19.0, 44.5] | 31.0 [22.0, 52.0] | 18.0 [10.0, 36.0] | 14.0 [9.0, 22.0] | 20.0 [11.0, 37.0] |
| COVID-19 severity |  |  |  |  |  |  |
| Fatal | 6 (1.5) | 89 (2.6) | 85 (1.6) | 6699 (17.6) | 594 (3.2) | 7473 (11.4) |
| Critical | 27 (6.7) | 129 (3.8) | 190 (3.5) | 973 (2.6) | 235 (1.3) | 1554 (2.4) |
| Severe | 29 (7.2) | 456 (13.4) | 1046 (19.2) | 11335 (29.8) | 3908 (21.1) | 16774 (25.5) |
| Mild to moderate | 343 (84.7) | 2728 (80.2) | 4127 (75.8) | 18984 (50.0) | 13827 (74.5) | 40009 (60.8) |
| **Microbiological cultures** |  |  |  |  |  |  |
| Blood culture | 98 (24.2) | 592 (17.4) | 640 (11.7) | 7733 (20.4) | 3665 (19.7) | 12728 (19.3) |
| Sputum culture | 31 (7.7) | 199 (5.8) | 251 (4.6) | 2246 (5.9) | 1212 (6.5) | 3939 (6.0) |
| Urine culture | 31 (7.7) | 238 (7.0) | 254 (4.7) | 6049 (15.9) | 3028 (16.3) | 9600 (14.6) |
| **Baseline drugs and COVID-19 vaccination** |  |  |  |  |  |  |
| COVID-19 vaccine doses, number<sup>b</sup> |  |  |  |  |  |  |
| 0 | 405 (100.0) | 3402 (100.0) | 5448 (100.0) | 20418 (53.7) | 4031 (21.7) | 33704 (51.2) |
| 1 | NA | NA | 0 (0.0) | 6710 (17.7) | 568 (3.1) | 7278 (11.1) |
| 2 | NA | NA | 0 (0.0) | 9236 (24.3) | 5100 (27.5) | 14336 (21.8) |
| 3 | NA | NA | 0 (0.0) | 1623 (4.3) | 8000 (43.1) | 9623 (14.6) |
| 4 | NA | NA | 0 (0.0) | 4 (0.0) | 865 (4.7) | 869 (1.3) |
| Antibacterial drugs | 4 (1.0) | 103 (3.0) | 127 (2.3) | 7170 (18.9) | 1839 (9.9) | 9243 (14.0) |
| Respiratory drugs | 1 (0.2) | 44 (1.3) | 53 (1.0) | 4023 (10.6) | 1038 (5.6) | 5159 (7.8) |
| Corticosteroids | 0 (0.0) | 34 (1.0) | 26 (0.5) | 4133 (10.9) | 923 (5.0) | 5116 (7.8) |
| Coronavirus antivirals | 0 (0.0) | 0 (0.0) | 0 (0.0) | 1865 (4.9) | 1613 (8.7) | 3478 (5.3) |
| **Baseline medical conditions** |  |  |  |  |  |  |
| Hypertension | 41 (10.1) | 712 (20.9) | 878 (16.1) | 21176 (55.7) | 8256 (44.5) | 31063 (47.2) |
| Diabetes mellitus | 23 (5.7) | 390 (11.5) | 486 (8.9) | 10612 (27.9) | 4048 (21.8) | 15559 (23.6) |
| Cardiovascular disease | 6 (1.5) | 180 (5.3) | 173 (3.2) | 9169 (24.1) | 3058 (16.5) | 12586 (19.1) |
| Malignancy | 4 (1.0) | 45 (1.3) | 41 (0.8) | 2487 (6.5) | 950 (5.1) | 3527 (5.4) |
| Chronic kidney disease | 0 (0.0) | 21 (0.6) | 16 (0.3) | 2781 (7.3) | 605 (3.3) | 3423 (5.2) |
| Chronic obstructive pulmonary disease | 0 (0.0) | 29 (0.9) | 21 (0.4) | 2112 (5.6) | 556 (3.0) | 2718 (4.1) |
| Chronic liver disease | 2 (0.5) | 35 (1.0) | 50 (0.9) | 1185 (3.1) | 389 (2.1) | 1661 (2.5) |
| Asthma | 0 (0.0) | 21 (0.6) | 33 (0.6) | 574 (1.5) | 267 (1.4) | 895 (1.4) |
| Rheumatic disease | 0 (0.0) | 2 (0.1) | 7 (0.1) | 252 (0.7) | 116 (0.6) | 377 (0.6) |
| Neutropenia | 0 (0.0) | 3 (0.1) | 1 (0.0) | 157 (0.4) | 67 (0.4) | 228 (0.3) |
| Organ transplant | 0 (0.0) | 3 (0.1) | 1 (0.0) | 157 (0.4) | 58 (0.3) | 219 (0.3) |

<sup>a</sup>Distribution of overall patient days for those admitted to an intensive care unit/high-dependency unit.
<sup>b</sup>COVID-19 vaccines became available locally beginning in February 2021 (i.e., wave 4).
Abbreviations: COVID-19, coronavirus disease 2019; F, female; ICU, intensive care unit; IQR, interquartile range; M, male; NA, not applicable.
Variables are shown as number of patients (%) unless otherwise indicated. Epidemic waves were defined as starting on the following dates: 1 January 2020 (waves 1-2), 1 July 2020 (wave 3), 1 November 2020 (wave 4), 31 December 2021 (wave 5E), and 23 May 2022 (wave 5L).

The number and characteristics of patients admitted to hospital varied by epidemic wave (**Table 1**). Patients admitted prior to wave 5 were younger, had fewer comorbidities, and more often had milder infections. The median number of days present in hospital declined over time as criteria for hospital discharge evolved and hospital capacity became limited during wave 5E. More than half of the patients admitted during wave 5E had not received any doses of COVID-19 vaccine especially among the 15,877 patients aged over 80 years (63.1% of these patients were not vaccinated). However, by wave 5L 78.3% of patients had received at least one dose of COVID-19 vaccine.

### Antibacterial drug prescriptions

During their hospital admission, 35,507 (54.0%) patients were prescribed an antibacterial drug, with the lowest prevalence in wave 4 (28.0%) and highest in wave 5E (64.6%; **Table 2**). Among those receiving an antibacterial drug, most were prescribed on the day of admission (69.5%) or within 2 days of admission (89.8%). Few young people < 20 years received an antibacterial drug prescription (n=747, 2.1%). In contrast, adults ≥ 80 years accounted for nearly half the patients receiving antibacterial treatment. The overall prevalence of antibacterial drug use by baseline characteristic and disease severity is presented in **Supplementary Table 3**. Nearly all patients admitted to ICU or with critical or fatal disease severity were prescribed an antibacterial drug.

**Table 2.**
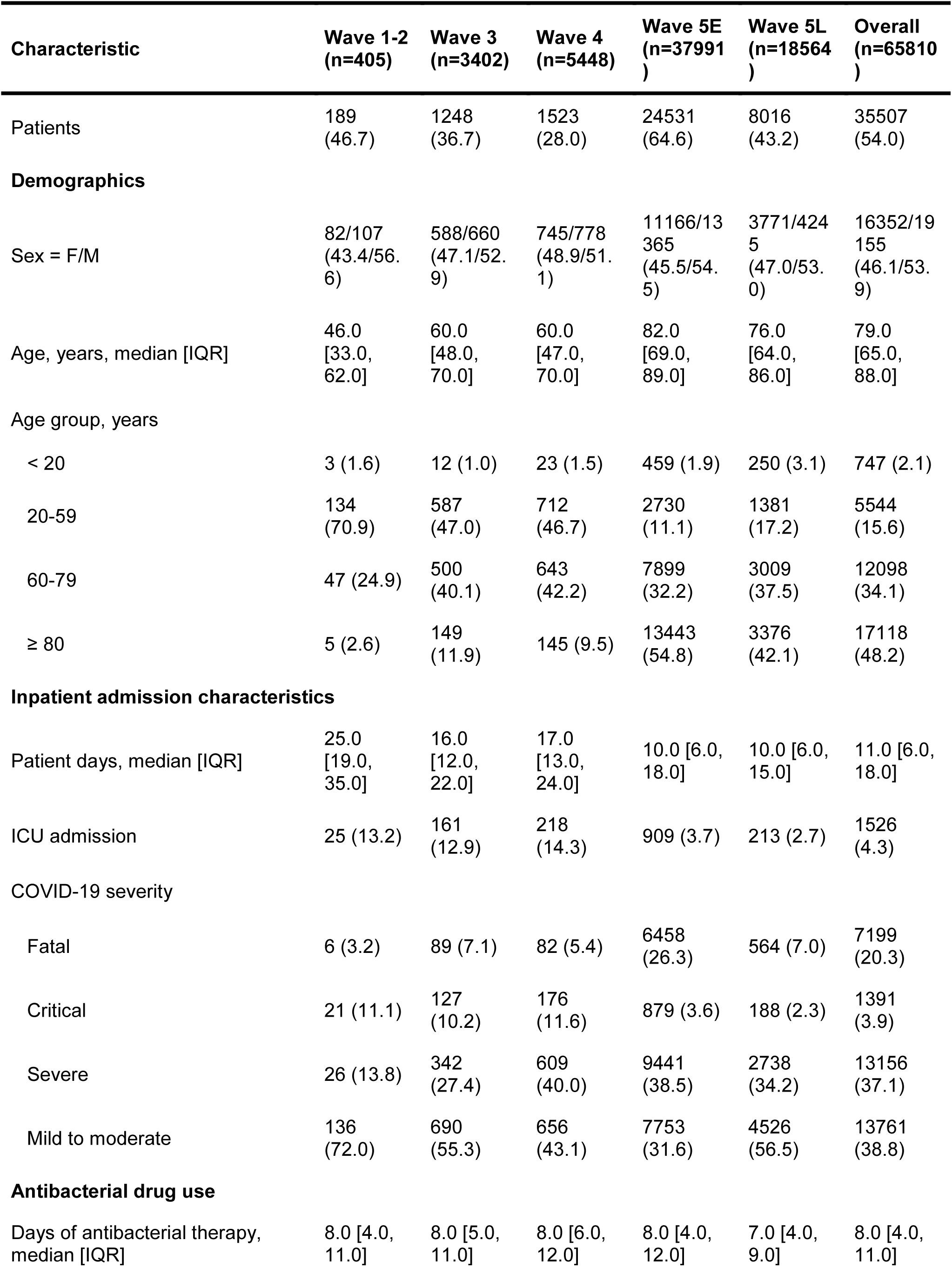
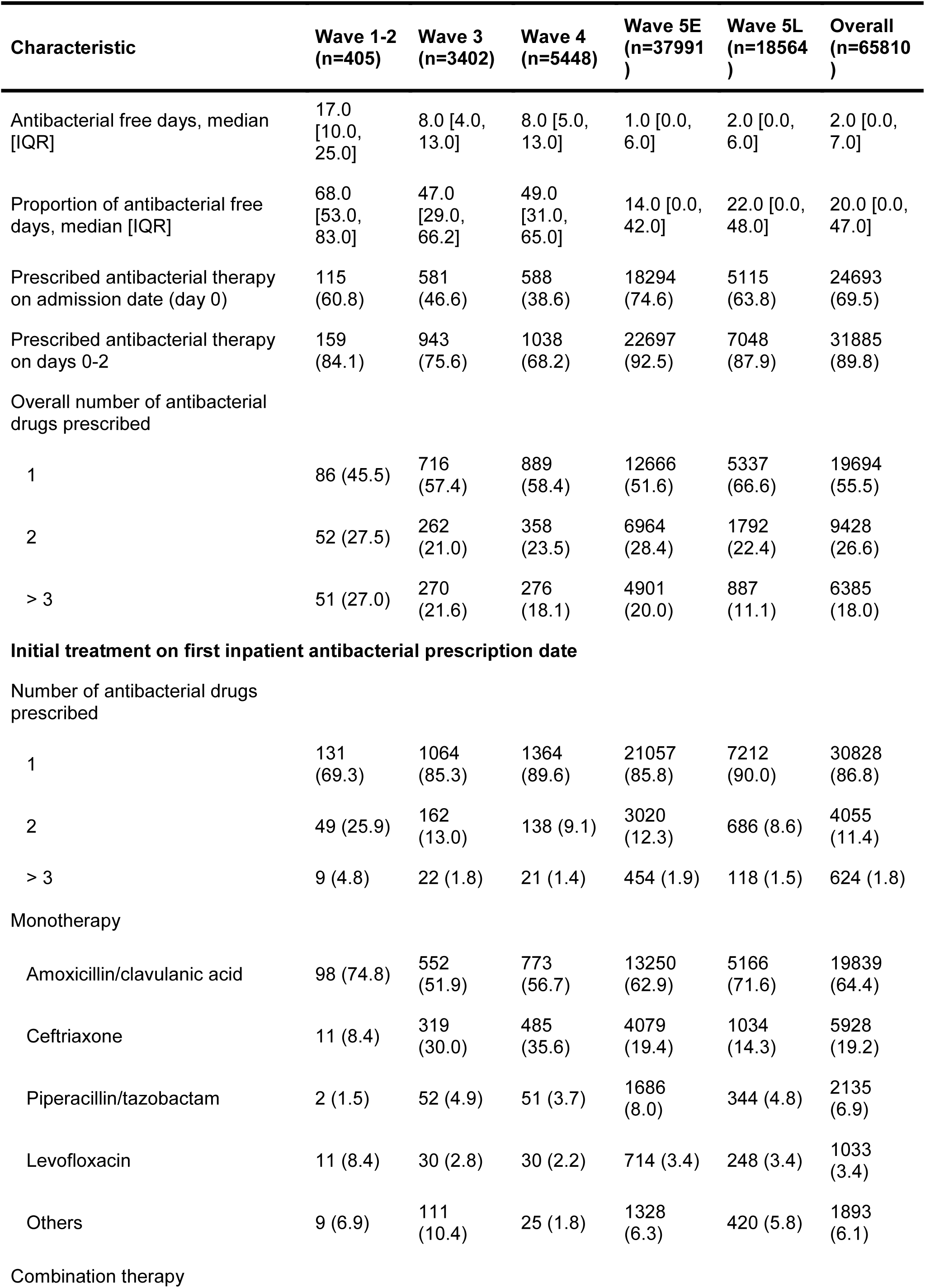
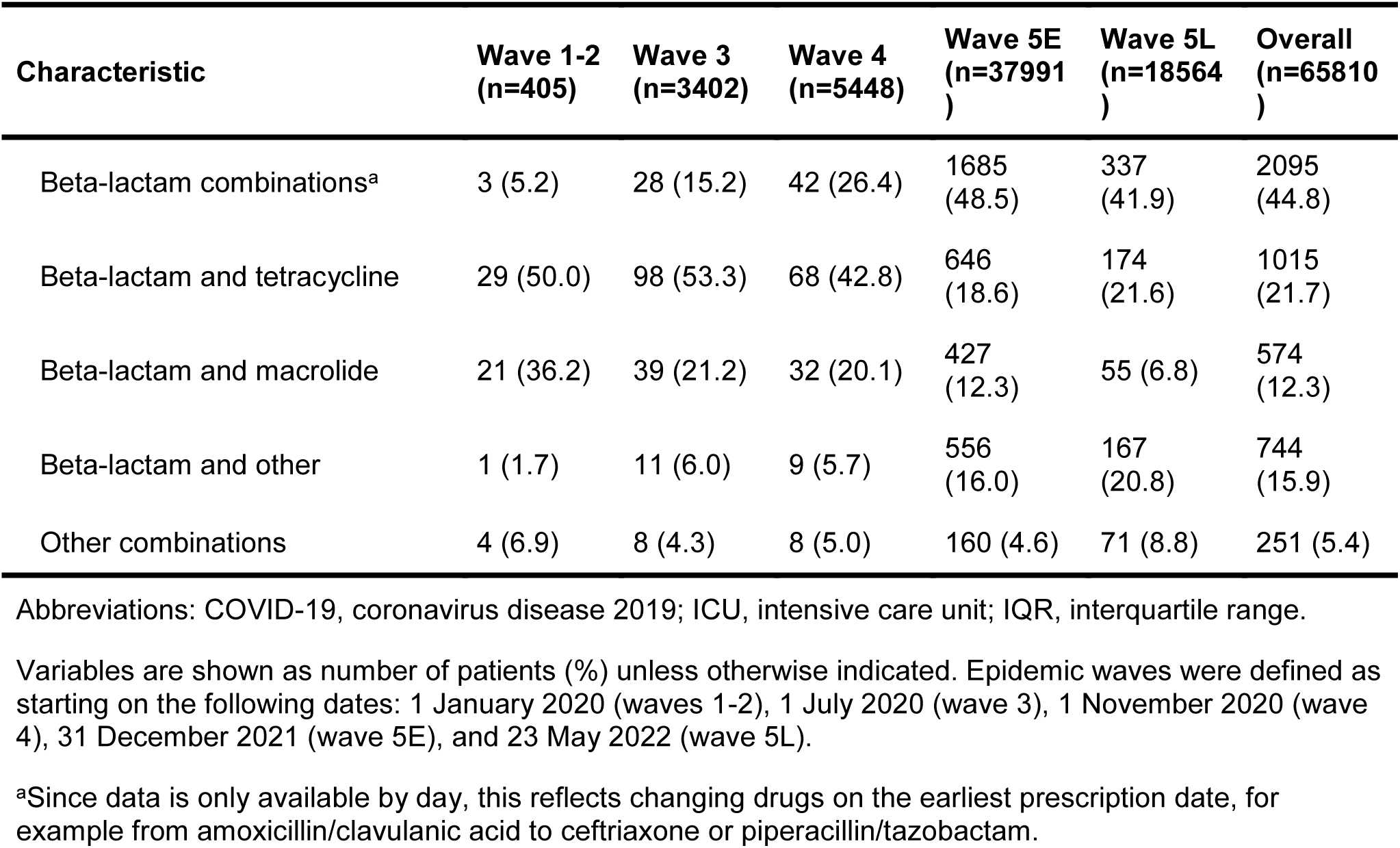
Characteristics of inpatients prescribed an antibacterial drug stratified by epidemic wave

Throughout the pandemic, the weekly use of antibacterial drugs in COVID-19 cases measured by DOT closely tracked hospital admissions for COVID-19 with substantially higher prescriptions observed during wave 5E although smaller variations in weekly prescribing rates (measured as DOT/1000 PD) occurred across waves (**Figure 1**). Antibacterial drugs were prescribed at an overall rate of 550.5 DOT/1000 PD (**Supplementary Table 4**). The most commonly used antibacterial drug classes were broad-spectrum penicillins (BNF 5.1.1.3), antipseudomonal penicillins (BNF 5.1.1.4), and cephalosporins (BNF 5.1.2.1) (**Figure 1C**). Overall, rates of use ranged from 246.9 DOT/1000 PD in waves 1-2 to 661.2 DOT/1000 PD in wave 5E (**Figure 1C** and **Supplementary Figure 3**). There was limited prescribing of macrolides. Tetracyclines were prescribed for COVID-19 patients, more often in waves 1-2, and higher prescriptions rates for carbapenems and antipseudomonal penicillins were observed in wave 5E than in earlier waves and wave 5L (**Figure 1C**).

Amoxicillin/clavulanic acid, piperacillin/tazobactam, and ceftriaxone were the three most prescribed antibacterial drugs and together accounted for 67.9% of the total antibacterial DOT prescribed to patients with COVID-19 (**Supplementary Table 5**). Throughout each epidemic wave, use of the most commonly prescribed antibacterial drugs was consistently higher among COVID-19 patients with fatal and critical disease severity than in patients with mild to moderate disease (**Supplementary Figure 4**). Amoxicillin/clavulanic acid, ceftriaxone, azithromycin, amoxicillin, cefotaxime, and doxycycline were frequently initiated within two days of admission, while drugs that are typically used to treat hospital-acquired bacterial infections, such as piperacillin/tazobactam, meropenem, ceftazidme, vancomycin, and linezolid, were initiated later (**Supplementary Table 5**).

### AWaReness of antibacterial drug prescribing

The majority of the antibacterial drugs prescribed to patients with COVID-19 during the study period belonged to the groups of access (47.8% of DOT) and watch (49.5%), and the rest were reserve (0.9%) and not recommended (1.8%) antibacterial drugs.

Rates of access and watch drug use increased rapidly in wave 5E and stabilized thereafter, and there were punctuated periods of greater use of reserve antibacterial drugs during waves 1-2, 3, and 4 (**Figure 2A-B**). Across the five defined epidemic waves, wave 5E had the highest prescribing rates of AWaRe drugs except for the reserve group, while in wave 5L the use of watch, reserve and not recommended drugs was comparable to or lower than in waves 1-4 in DOT/1000 PD, and considerably lower than all other waves when measured by DOT/1000 patients (**Supplementary Figure 5**). Amoxicillin/clavulanic acid, piperacillin/tazobactam, and minocycline (IV) were respectively the most prescribed access, watch, and reserve drugs and cefoperazone/sulbactam was the only not recommended group drug prescribed (**Figure 2C**). Weekly DOT showed that an increasing number of days treated with watch or reserve drugs appeared in more severe infections, which was largely consistent across different epidemic waves (**Supplementary Figure 6**).

**Figure 2.**
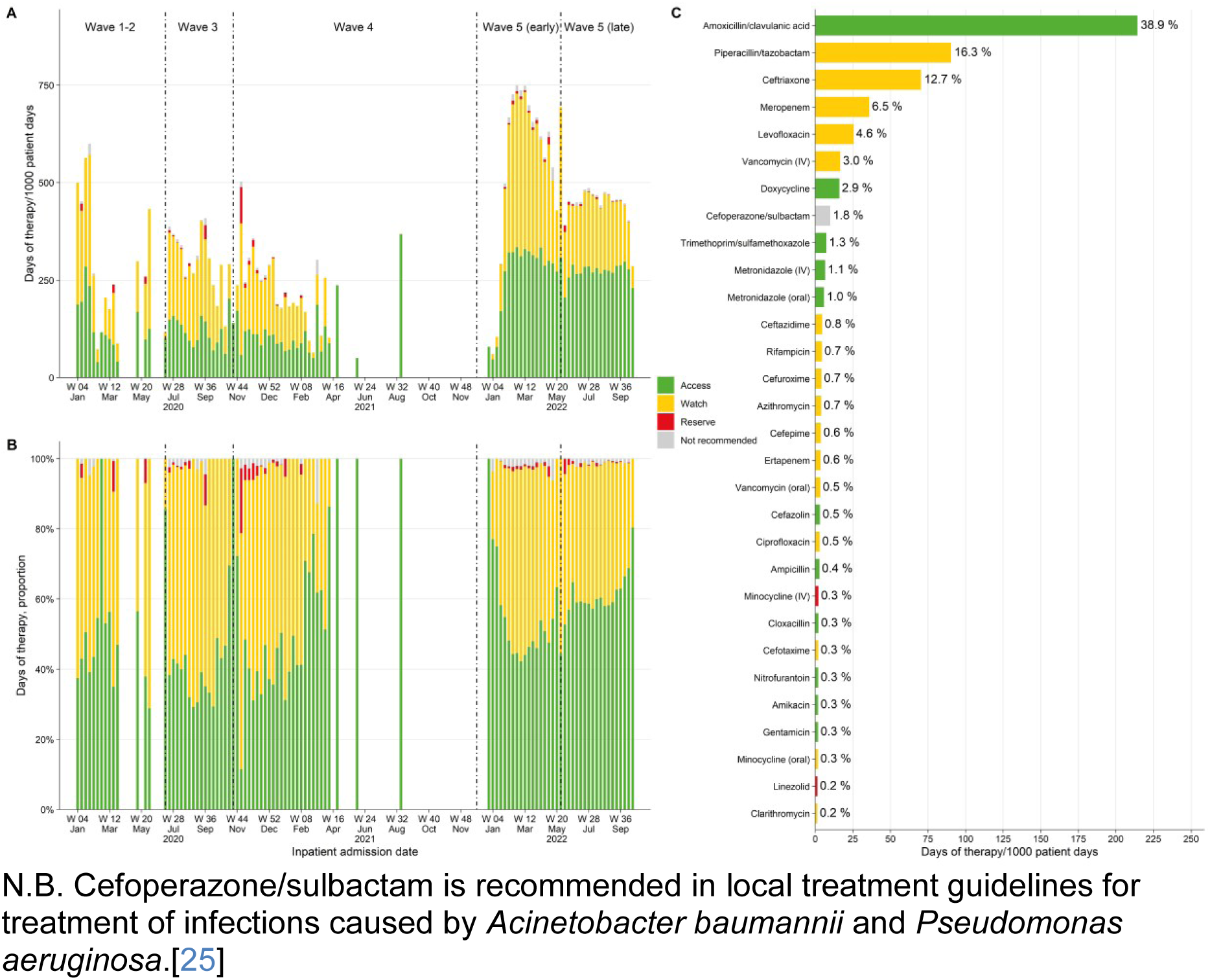
Weekly rates of antibacterial drug consumption (**A**), proportion of antibacterial days of therapy (**B**), and the 30 most prescribed antibacterial drugs (**C**) classified according to WHO AWaRe group. For each drug in **C**, the label on the right indicates the drug’s share of all antibacterial drug days of therapy. N.B. Cefoperazone/sulbactam is recommended in local treatment guidelines for treatment of infections caused by *Acinetobacter baumannii* and *Pseudomonas aeruginosa*.[25]

### Factors associated with antibacterial drug prescription

Factors associated with an increased odds of an inpatient antibacterial drug prescription included age ≥ 60 years (vs. 20-59 years); living in a residential care home for the elderly (vs. not a care home resident); fatal or critical COVID-19 (vs. severe COVID-19); and presence of the following factors during the pre-defined baseline period, such as blood, sputum, or urine culture, a prescription for an antibacterial drug, corticosteroid, or respiratory drug, and a diagnosis of chronic obstructive pulmonary disease or neutropenia (**Figure 3**). There were weak or no associations for the other baseline medical conditions included in the model. In contrast, we observed a strong graded association for a reduced odds of an inpatient antibacterial prescription with more doses of COVID-19 vaccine received 14 days prior to admission (**Figure 3**). Furthermore, pre-admission treatment with coronavirus specific antivirals was also negatively associated with an antibacterial drug prescription.

**Figure 3.**
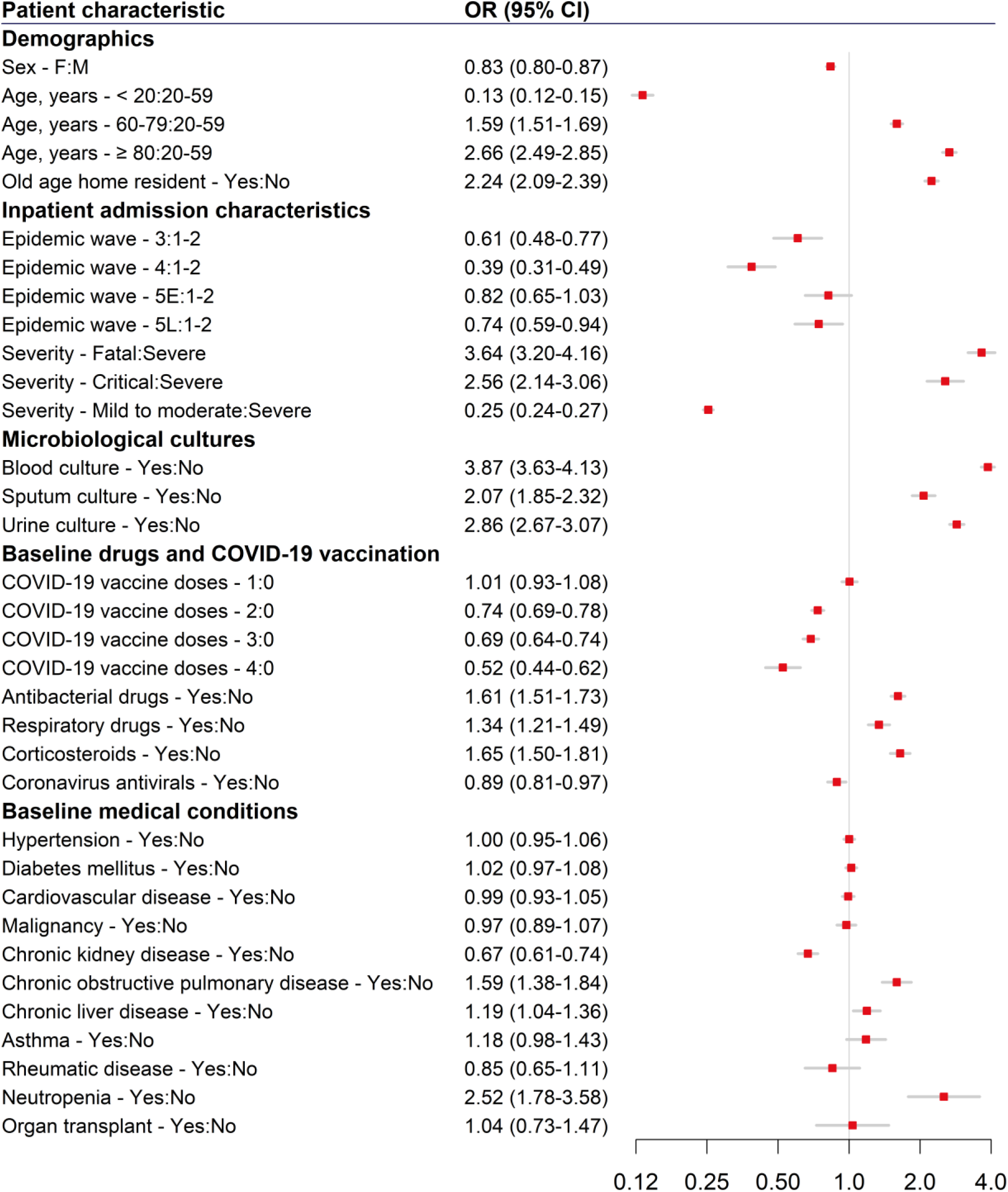
Association between baseline patient characteristics and disease severity with an inpatient antibacterial prescription

## DISCUSSION

This study is one of the largest analyses of inpatient prescribing of antibacterial drugs in adults and children admitted to hospital with COVID-19. Our findings demonstrate a high prescription rate of antibacterial drugs among hospitalized patients infected in the local community in Hong Kong during the pandemic, particularly in the largest wave caused by the Omicron BA.2 subvariants (wave 5E). Furthermore, we identified important variations in prescribing specific types of drugs among patients with different characteristics and highlighted the baseline factors associated with antibacterial prescriptions.

The high-quality patient data collected in Hong Kong in response to the COVID-19 pandemic allowed us to conduct a comprehensive examination of antibacterial drug prescribing throughout multiple epidemic waves. In a point-prevalence survey carried out in Scotland in April 2020, 45.0% of patients with COVID-19 were prescribed antibacterial drugs.[19] This estimate was similar to our estimate (46.7%) for waves 1-2. Despite evidence from a seminal systematic review demonstrating a low prevalence (∼15%) of bacterial co-infections or secondary infections among hospitalized patients with COVID-19 during the early days of the pandemic,[4] a substantial proportion of patients in our study were prescribed antibacterial drugs in later epidemic waves, revealing a clear mismatch between the prevalence of bacterial infections and antibacterial prescribing. This inconsistency has been reported in studies conducted worldwide. One of the largest studies of hospitalized patients with SARS-CoV-2 in the United Kingdom between February and June 2020 showed that 85.2% of admitted patients received an antibacterial drug prescription and that the prescribing trend was declining towards the end of the study period.[20] A scoping review of studies published until March 2021 also showed that the prescribing of antibacterial drugs declined between June 2020 to March 2021.[3] This aligns with the time trends observed in our study. However, a surge in antibacterial drug use occurred in early 2022 with the Omicron variant, a concerning finding that may have implications for the development of AMR in Hong Kong.

Our findings differ from those of studies conducted in other high-income Asian countries. A point prevalence survey of 577 hospitalized patients with COVID-19 in Singapore conducted in April 2020 reported that 6.2% of the patients were prescribed antibacterial drugs and the most prescribed drugs were amoxicillin/clavulanic acid (51% of prescriptions), clarithromycin, and piperacillin/tazobactam.[21] Half of the prescriptions for amoxicillin/clavulanic acid were judged inappropriate by the authors.[21] Using data from January 2020 to November 2021, a study of 66,912 hospitalized patients with COVID-19 in Japan found that 16.15% were prescribed an antibacterial drug and the most prescribed drugs were ceftriaxone, ampicillin/sulbactam, and azithromycin. Regional variations in antimicrobial prescribing among patients with similar infections are well documented.[22] These variations could be the result of the local epidemiology of drug resistance, local treatment guidelines, and prescribing culture. We identified infrequent use of macrolides but a higher overall prevalence of antibacterial drug prescribing compared to these two studies. Due to its potential antiviral and anti-inflammatory effects, azithromycin was the most frequently prescribed antibacterial drug, especially early in the pandemic, among patients with COVID-19.[3] On the contrary, we observed limited azithromycin prescribing in Hong Kong because local experts did not recommend its use as a treatment for COVID-19 due to uncertain evidence of efficacy. Furthermore, high levels of local resistance to macrolides among *Mycoplasma pneumoniae* and *Streptococcus pneumoniae* isolates have led to reductions over time in macrolide prescribing among inpatients.[23] In accordance with local treatment guidelines, clinicians prescribe more often levofloxacin or doxycycline as the preferred atypical organism coverage for community-acquired pneumonia.[24,25]

The pandemic response measures applied in Hong Kong required all confirmed COVID-19 cases to be isolated in designated facilities mainly isolation wards in public hospitals. This allowed us to examine antibacterial prescriptions together with recorded characteristics of the patients. By using data until October 2022, we were able to include information on baseline vaccination and oral antiviral drug treatment for patients admitted from early 2021 onward, with the majority of hospitalizations caused by the Omicron variants, which has not been addressed in previous studies. Our findings support that patients with ≥ 2 doses of COVID-19 vaccine and outpatient treatment with antiviral drugs prior to hospital admission had a relatively lower odds of receiving an inpatient antibacterial prescription. This indicates a lower risk of suspected bacterial co-infection or secondary infection among these patients or perhaps different prescribing practices for such patients.

By quantifying antimicrobial prescribing using standard metrics, we have demonstrated the feasibility of conducting surveillance on antibacterial use using routinely collected inpatient data. At global level, the WHO is undertaking a large-scale clinical platform to understand antibacterial drug prescribing in COVID-19,[26] evidence that could establish benchmarks for patients admitted with COVID-19 and other respiratory viral infections, and could serve to make comparisons among treatment patterns within health systems, countries, and regions. Our research complements and supports these global efforts by providing comprehensive data on the prevalence and types of antibacterial drugs used according to the AWaRe classification among a diverse cohort of inpatients from Hong Kong.

Nonetheless, some limitations exist in this study. The available data did not include prescription indications nor were we able to assess the appropriateness of antibacterial therapy. These limitations could be addressed in future studies that prospectively collect information on the appropriateness of therapy and data are needed to determine the occurrence of bacterial co-infection (concurrent with SARS-CoV-2 infection), super-infection, or secondary infection. In addition, the baseline data used for the analysis was collected from outpatient clinics operated by the HA and public hospitals, which might have missed information from patients who had consultation at private hospitals or clinics. However, public hospitals provide 80%-90% of hospital bed days in Hong Kong.

## Conclusions

Antibacterial drug prescribing varied substantially among inpatients with COVID-19 indicating an urgent need for the development of standardized and evidence-based diagnostic and antibacterial treatment guidelines. Ongoing surveillance of inpatient antibacterial drug prescribing can be used for targeting antimicrobial stewardship activities and should be implemented as a key component of healthcare systems in response to both COVID-19 and AMR.

## Data Availability

Patient data used in the analysis were derived from electronic health records managed by the Hospital Authority and other patient information collected by the Centre for Health Protection during case finding and contact tracing. Access to the underlying data used in this study is subject to the approval of the two agencies.

## ACKNOWLEDGMENTS

The authors thank the Hospital Authority and the Department of Health of the Government of Hong Kong Special Administrative Region for providing the data for the analysis, Ms Kitty TM Ng (antimicrobial stewardship pharmacist) from Queen Elizabeth Hospital, Hong Kong, for her comments on the manuscript, and Julie Au and Chloe Chui for their technical support.

## Contributions

Conceptualization: Joseph E Blais, Peng Wu

Resources and funding acquisition: Peng Wu, Benjamin J Cowling

Supervision: Peng Wu, Benjamin J Cowling

Writing, original draft: Joseph E Blais

Writing, review and editing: Weixin Zhang, Yun Lin, Peng Wu, Vincent CC Cheng, Celine SL Chui, Benjamin J Cowling

Formal analysis: Joseph E Blais

Data curation: Yun Lin

Visualization: Joseph E Blais

## SUPPLEMENTARY MATERIAL

**Supplementary Figure 1.**
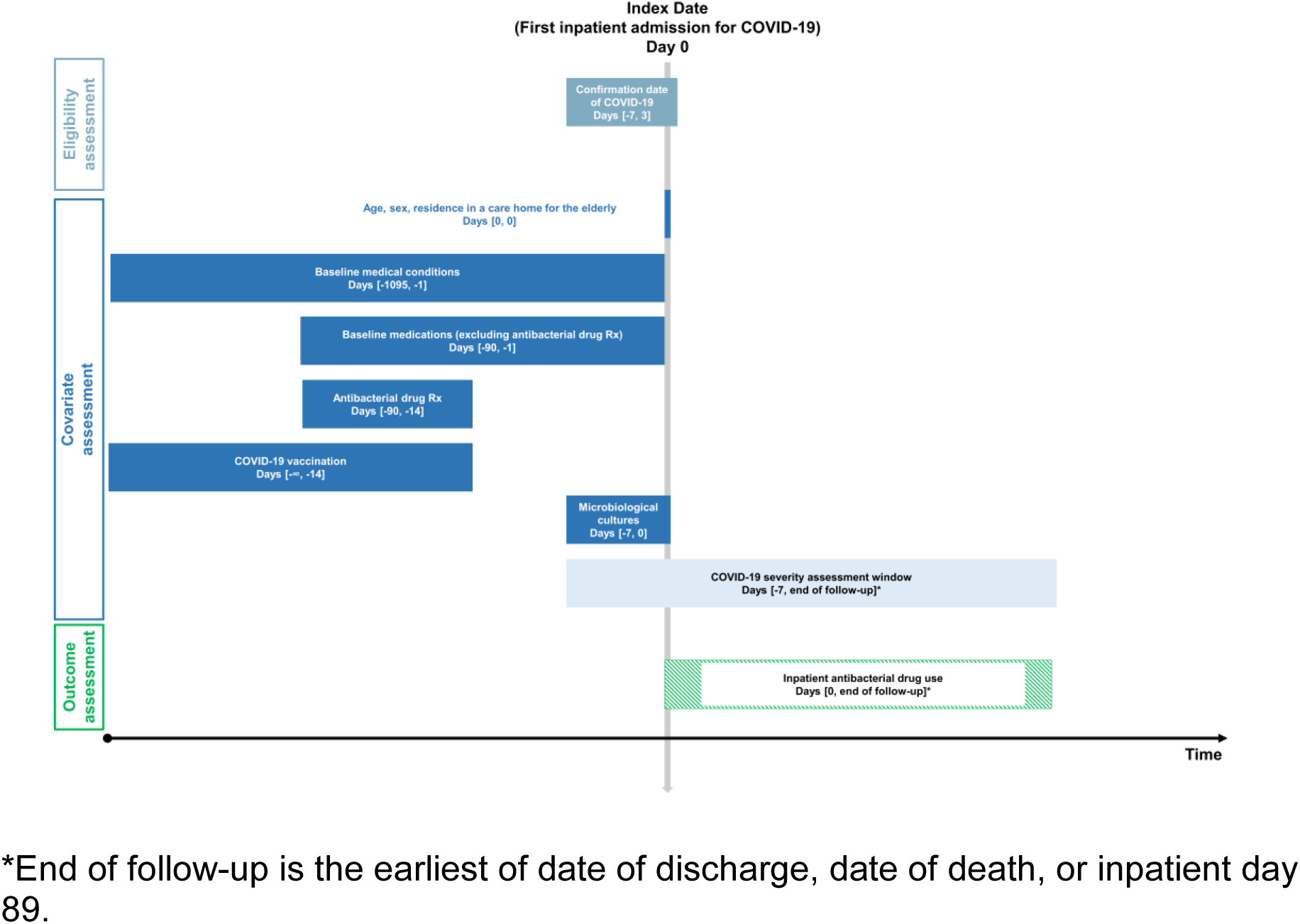
Overview of the study cohort design and assessment windows

**Supplementary Table 1.** Table of variable definitions used in the study

| Variable type | Variable name | Data Source | Definition | Window |
| --- | --- | --- | --- | --- |
| **Time anchor** | Date of hospital admission (index date or day 0) | Hospitalization records and discharge status | Defined as the earliest (first) hospital admission date with date of COVID-19 confirmation during the eligibility window. | [21 January 2020, 30 September 2022] |
| **Time anchor** | Date of hospital discharge | Hospitalization records and discharge status<br>Date of death | Defined as the earliest date of death or discharge. | [21 January 2020, 30 October 2022] |
| **Time anchor** | End of follow-up | Hospitalization records and discharge status<br>Date of death | Defined as the earliest date of discharge, death or inpatient day 89. | NA |
| **Time anchor** | Date of COVID-19 confirmation | CHP case data | Date of COVID-19 confirmation by the CHP. | [-7, 3] |
| **Time anchor** | Epidemic wave | Hospitalization records | Date of hospital admission categorized into corresponding epidemic waves. | **Wave 1-2:** 1 January 2022 to 30 June 2020<br>**Wave 3:** 1 July 2022 to 31 October 2020<br>**Wave 4:** 1 November 2020 to 30 December 2021<br>**Wave 5E (early):** 31 December 2021 to 22 May 2022<br>**Wave 5L (late):** 23 May 2022 onward |
| **Person time** | Patient days | Hospitalization records and discharge status | Defined as date of hospital discharge – index date + 1 day. Censored at the end of follow-up. | [0, end of follow-up] |
| **COVID-19 case classification** | Case classification | CHP case data | Cases classified by as either local or imported.<br>**Local cases** are defined as:<br>local, close contact of local, locally acquired case,<br>possibly local, close contact of possibly local and missing.<br>**Imported cases** are defined as:<br>Imported and close contact of imported. | NA |
| Demographics | Age | Demographics | As reported by CHP or HA, in years. | [0, 0] |
| Demographics | Sex | Demographics | Male or female. | [0, 0] |
| Demographics | Residence in a care home for the elderly (Yes/No) | Inpatient | Admitted from a residential care home for the elderly. | [0, 0] |
| Clinical outcomes | ICU Admission (Yes/No) | Inpatient transactions | Any admission record to intensive care unit/high-dependency unit. | [0, end of follow-up] |
| Clinical outcomes | In-hospital death (Yes/No) | Inpatient transactions<br>Date of death | Discharge destination code is "D" – Death<br>**OR**<br>If date of death is available, date of death is on or before the discharge date. | [0, discharge date] |
| Baseline medical conditions | Cardiovascular disease (Yes/No) | Inpatient diagnosis<br>General outpatient diagnosis | A diagnosis of any of the following cardiovascular diseases defined as the following diagnosis codes (including all subcodes and codes in any position).<br>**Ischemic heart disease**<br>ICD-9-CM code: 410-414, V45.81<br>**OR**<br>ICPC-2 code: K74, K75, K76<br>**Cerebrovascular disease**<br>ICD-9-CM code: 430-438<br>**OR**<br>ICPC-2 code: K89, K90, K91<br>**Peripheral vascular disease**<br>ICD-9-CM code: 440-443, 447.1, 557.1, 557.9, V43.4<br>**OR**<br>ICPC-2 code: K92<br>**Heart failure**<br>ICD-9-CM code: 398.91, 402.01, 402.11, 402.91, 404.01, 404.03, 404.11, 404.13, 404.91, 404.93, 428.x, 425.4, | [-1095, -1] |
|  |  |  | 425.5, 425.7, 425.8,<br>425.9, 428<br>**OR**<br>ICPC-2 code: K77 |  |
| **Baseline medical conditions** | Chronic kidney disease (Yes/No) | Inpatient diagnosis<br>General outpatient diagnosis | A diagnosis of chronic kidney disease defined as ICD-9-CM code (including all subcodes and codes in any position): 403.01, 403.11, 403.91, 404.02, 404.03, 404.12, 404.13, 404.92, 404.93, 582, 583.0-583.7, 585, 586, 588.0, V42.0, V45.1, V56 | [-1095, -1] |
| **Baseline medical conditions** | Hypertension (Yes/No) | Inpatient diagnosis<br>General outpatient diagnosis<br>Prescriptions | A diagnosis of hypertension defined as ICD-9-CM code (including all subcodes and codes in any position): 401-405<br>**OR**<br>ICPC-2 code: K86, K87<br>**OR**<br>A prescription dispensing date for any antihypertensive medication included in BNF Sections:<br>2.5.4 Alpha-adrenoreceptor blocking drugs<br>2.2.1 Thiazides and related diuretics<br>2.2.3 Potassium-sparing diuretics and aldosterone antagonists<br>2.2.4 Potassium-sparing diuretics with other diuretics<br>2.4 Beta-adrenoceptor blocking drugs<br>2.5.1 Vasodilator antihypertensive drugs<br>2.5.2 Centrally acting antihypertensive drugs<br>2.6.2 Calcium-channel blockers<br>2.5.5.1 Angiotensin-converting enzyme inhibitors<br>2.5.5.2 Angiotensin-II receptor antagonists | [-1095, -1]<br>[-90, -1] |
|  |  |  | 2.5.5.3 Renin inhibitors |  |
| **Baseline medical conditions** | Diabetes mellitus (Yes/No) | Inpatient diagnosis<br>General outpatient diagnosis<br>Prescriptions | A diagnosis of diabetes mellitus defined as ICD-9-CM code (including all subcodes and codes in any position): 250, 790.2<br>OR<br>ICPC-2 code: T89, T90<br>OR<br>A prescription dispensing date for any antidiabetic medication included in BNF Sections:<br>6.1.1.1 Short-acting insulins<br>6.1.1.2 Intermediate- and long-acting insulins<br>6.1.2.1 Sulphonylureas<br>6.1.2.2 Biguanides<br>6.1.2.3 Other antidiabetic drugs | [-1095, -1]<br>[-90, -1] |
| **Baseline medical conditions** | Chronic obstructive pulmonary disease (Yes/No) | Inpatient diagnosis<br>General outpatient diagnosis | A diagnosis of COPD defined as ICD-9-CM code (including all subcodes and codes in any position): 490-492, 494-6, 500-505, 506.4, 508.1, 508.8<br>OR<br>ICPC-2 code: R95 | [-1095, -1] |
| **Baseline medical conditions** | Asthma (Yes/No) | Inpatient diagnosis<br>General outpatient diagnosis | A diagnosis of asthma defined as ICD-9-CM code (including all subcodes and codes in any position): 493<br>OR<br>ICPC-2 code: R96 | [-1095, -1] |
| **Baseline medical conditions** | Chronic liver disease (Yes/No) | Inpatient diagnosis<br>General outpatient diagnosis | A diagnosis of liver disease defined as ICD-9-CM code (including all subcodes and codes in any position):<br>571, 572.3, 572.4, 572.8, 573.0, 573.1, 573.2, 573.4, 573.5, 573.8, 573.9<br>OR<br>ICPC-2 code: D97 | [-1095, -1] |
| **Baseline medical conditions** | Rheumatic disease and related conditions (Yes/No) | Inpatient diagnosis<br>General outpatient diagnosis | A diagnosis of rheumatoid arthritis and related conditions defined as ICD-9-CM code (including all<br>subcodes and codes in any position): 446.5, 710.0-710.9, 714.0-714.2, 714.8, 725<br>**OR**<br>ICPC-2 code: L88 | [-1095, -1] |
| **Baseline medical conditions** | Malignancy (Yes/No) | Inpatient diagnosis<br>General outpatient diagnosis | Diagnosis of malignancy defined as ICD-9-CM code (including all subcodes and codes in any position): 140-149, 150-159, 160-165, 170-176, 179-189, 190-199, 200-208<br>**OR**<br>ICPC-2 code: B72-B74, D74-D77, L71, N74, R84, R85, S77, T71, U75- U77, X75- X77, Y77, Y78 | [-1095, -1] |
| **Baseline medical conditions** | Organ transplant (Yes/No) | Inpatient diagnosis | Diagnosis of solid organ transplant (kidney, heart, lung, or liver) defined as ICD-9-CM code (including all subcodes and codes in any position): V42.0, V42.1, V42.6, V42.7 | [-1095, -1] |
| **Baseline medical conditions** | Neutropenia (Yes/No) | Inpatient diagnosis | Diagnosis of neutropenia defined as ICD-9-CM code (including all subcodes and codes in any position): 288.0 | [-1095, -1] |
| **Baseline drugs** | Respiratory drugs (Yes/No) | Prescriptions | A prescription dispensing date for any of the medications included in BNF Sections:<br>3.1.1.1 Adrenoceptor agonists<br>3.1.2 Antimuscarinic bronchodilators<br>3.1.3 Theophylline<br>3.1.4 Compound bronchodilator preparations<br>3.2 Corticosteroids (respiratory)<br>3.3.1 Cromoglycate and related therapy<br>3.3.2 Leukotriene receptor antagonists<br>3.3.3 Phosphodiesterase Type-4 inhibitors | [-90, -1] |
| Baseline drugs | Corticosteroids (Yes/No) | Prescriptions | A prescription dispensing date for any of the medications included in BNF Sections:<br>1.5.2 Corticosteroids<br>6.3.1 Replacement therapy<br>6.3.2 Glucocorticoid therapy<br>10.1.2.2 Corticosteroids | [-90, -1] |
| Baseline drugs | Antibacterial drugs (Yes/No) | Prescriptions | A prescription dispensing date for any of the medications included in BNF Section 5.1 Antibacterial drugs:<br>5.1.1.1 Benzylpenicillin and phenoxymethylpenicillin<br>5.1.1.2 Penicillinase-resistant penicillins<br>5.1.1.3 Broad-spectrum penicillins<br>5.1.1.4 Antipseudomonal penicillins<br>5.1.2.1 Cephalosporins<br>5.1.2.2 Carbapenems<br>5.1.2.3 Other beta-lactam antibiotics<br>5.1.3 Tetracyclines<br>5.1.4 Aminoglycosides<br>5.1.5 Macrolides<br>5.1.6 Clindamycin and lincomycin<br>5.1.7 Some other antibacterials<br>5.1.8 Sulfonamides and trimethoprim<br>5.1.9 Antituberculosis drugs<br>5.1.10 Antileprotic drugs<br>5.1.11 Metronidazole and tinidazole<br>5.1.12 Quinolones<br>5.1.13 Urinary-tract infections | [-90, -14] |
| Baseline drugs | Coronavirus specific antivirals (Yes/No) | Prescriptions | A prescription dispensing date for any of the medications, defined as drug item codes:<br>**remdesivir** (REMD01, REMD02 and REMD03)<br>**OR**<br>**paxlovid** (PAXL01 and | [-90, -1] |
|  |  |  | PAXL02)<br>**OR**<br>**molnupiravir**<br>(MOLN01) |  |
| **COVID-19 vaccination** | Number of COVID-19 vaccine doses (0 to 4) | Vaccination records | Total number of COVID-19 vaccine doses administered during the window. | [start of data, -14] |
| **Admission microbiological culture** | Blood culture (Yes/No) | Laboratory | Test date during the window and a test description of any of the following:<br>"Blood Culture", "Blood culture:", "Bactec culture and ST", "Blood culture :-", "Blood culture :", "Blood Culture :-" | [-7, 0] |
| **Admission microbiological culture** | Sputum culture (Yes/No) | Laboratory | Test date during the window and a test description of any of the following:<br>"Sputum Culture :", "Sputum Culture", "Sputum Culture :-", "Sputum culture :", "Respiratory Culture", "Microbiological test of respiratory specimens" | [-7, 0] |
| **Admission microbiological culture** | Urine culture (Yes/No) | Laboratory | Test date during the window and a test description of any of the following:<br>"Urine culture :", "Urine culture", "Urine Culture", "Urine Culture :-", "Microbiological test of urine specimens" | [-7, 0] |

**Abbreviations:** CHP, Centre for Health Protection; COVID-19, coronavirus disease 2019; HA, Hospital Authority; ICD-9-CM, International Classification of Diseases, Ninth Revision, Clinical Modification; ICPC-2, International Classification of Primary Care, 2nd edition; ICU, intensive care unit.

**Supplementary Table 2.** COVID-19 severity definitions used in the study. Day zero is defined as the date of hospital admission

| Severity | Definition | Window |
| --- | --- | --- |
| **Fatal** | In-hospital death | [0, date of discharge] |
| **Critical** | Any inpatient condition recorded as 'critical'*<br>**OR**<br>inpatient procedures for mechanical ventilation, intubation, or ECMO<br>**OR**<br>had any inpatient day present in ICU/HDU. | [0, end of follow-up] |
| **Severe** | Prescription dispensing date for remdesivir, baricitinib, tocilizumab, or dexamethasone (in BNF Section 6.3.2 and oral or injectable route)<br>**OR**<br>any oxygen saturation test result $\leq 90\%$<br>**OR**<br>any inpatient condition recorded as 'serious'*. | [0, earliest of day 9 or end of follow-up]<br>[-7, end of follow-up]<br>[0, end of follow-up] |
| **Mild to moderate** | Not meeting the definition of either fatal, critical or severe disease. | [0, end of follow-up] |

\* Inpatient condition as defined by the Hospital Authority:
- "Critical" – Patient in Intensive Care Unit, **OR** intubated, **OR** requires Extracorporeal Membrane Oxygenation (ECMO) **OR** in shock. - "Serious" – Patient requires oxygen supplement of 3 liters per minute or more.

**Supplementary Figure 2.**
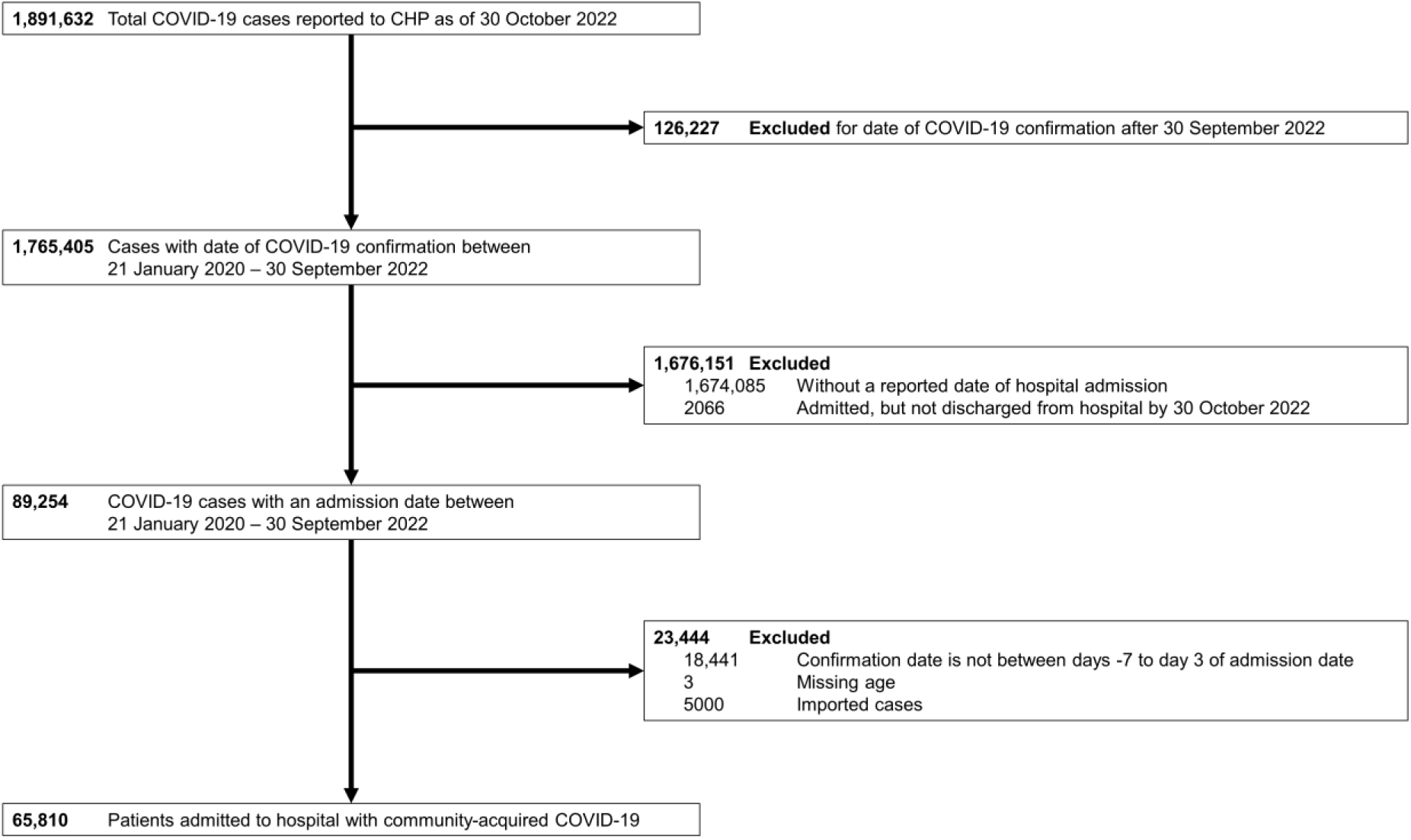
Flow diagram of patient inclusion in the study cohort

**Supplementary Table 3.** Prevalence (%) of antibacterial drug prescription by patient characteristic and stratified by epidemic wave. See Supplementary Table 1 for detailed definitions of each variable.

| Wave | 1-2 |  |  | 3 |  |  | 4 |  |  | 5E |  |  | 5L |  |  | Overall |  |  |
| --- | --- | --- | --- | --- | --- | --- | --- | --- | --- | --- | --- | --- | --- | --- | --- | --- | --- | --- |
|  | No.<br>with<br>Rx | No.<br>Total | % | No.<br>with<br>Rx | No.<br>Total | % | No.<br>with<br>Rx | No.<br>Total | % | No.<br>with<br>Rx | No.<br>Total | % | No.<br>with<br>Rx | No.<br>Total | % | No.<br>with<br>Rx | No.<br>Total | % |
| All patients | 189 | 405 | 46.7 | 1,248 | 3,402 | 36.7 | 1,523 | 5,448 | 28.0 | 24,531 | 37,991 | 64.6 | 8,016 | 18,564 | 43.2 | 35,507 | 65,810 | 54.0 |
| **Sex** |  |  |  |  |  |  |  |  |  |  |  |  |  |  |  |  |  |  |
| F | 82 | 191 | 42.9 | 588 | 1,765 | 33.3 | 745 | 2,861 | 26.0 | 11,166 | 18,110 | 61.7 | 3,771 | 8,967 | 42.1 | 16,352 | 31,894 | 51.3 |
| M | 107 | 214 | 50.0 | 660 | 1,637 | 40.3 | 778 | 2,587 | 30.1 | 13,365 | 19,881 | 67.2 | 4,245 | 9,597 | 44.2 | 19,155 | 33,916 | 56.5 |
| **Age group, years** |  |  |  |  |  |  |  |  |  |  |  |  |  |  |  |  |  |  |
| < 20 | 3 | 12 | 25.0 | 12 | 281 | 4.3 | 23 | 508 | 4.5 | 459 | 3,228 | 14.2 | 250 | 3,730 | 6.7 | 747 | 7,759 | 9.6 |
| 20-59 | 134 | 324 | 41.4 | 587 | 2,042 | 28.7 | 712 | 3,407 | 20.9 | 2,730 | 7,124 | 38.3 | 1,381 | 3,243 | 42.6 | 5,544 | 16,140 | 34.3 |
| 60-79 | 47 | 62 | 75.8 | 500 | 892 | 56.1 | 643 | 1,328 | 48.4 | 7,899 | 11,762 | 67.2 | 3,009 | 6,435 | 46.8 | 12,098 | 20,479 | 59.1 |
| ≥ 80 | 5 | 7 | 71.4 | 149 | 187 | 79.7 | 145 | 205 | 70.7 | 13,443 | 15,877 | 84.7 | 3,376 | 5,156 | 65.5 | 17,118 | 21,432 | 79.9 |
| **Old age home resident** |  |  |  |  |  |  |  |  |  |  |  |  |  |  |  |  |  |  |
| No | 189 | 405 | 46.7 | 1,248 | 3,402 | 36.7 | 1,523 | 5,448 | 28.0 | 13,948 | 26,005 | 53.6 | 7,072 | 17,290 | 40.9 | 23,980 | 52,550 | 45.6 |
|  | No. with Rx | No. Total | % | No. with Rx | No. Total | % | No. with Rx | No. Total | % | No. with Rx | No. Total | % | No. with Rx | No. Total | % | No. with Rx | No. Total | % |
| Yes | 0 | 0 | 0.0 | 0 | 0 | 0.0 | 0 | 0 | 0.0 | 10,583 | 11,986 | 88.3 | 944 | 1,274 | 74.1 | 11,527 | 13,260 | 86.9 |
| **COVID-19 severity<sup>a</sup>** |  |  |  |  |  |  |  |  |  |  |  |  |  |  |  |  |  |  |
| Fatal | 6 | 6 | 100.0 | 89 | 89 | 100.0 | 82 | 85 | 96.5 | 6,458 | 6,699 | 96.4 | 564 | 594 | 94.9 | 7,199 | 7,473 | 96.3 |
| Critical | 21 | 27 | 77.8 | 127 | 129 | 98.4 | 176 | 190 | 92.6 | 879 | 973 | 90.3 | 188 | 235 | 80.0 | 1,391 | 1,554 | 89.5 |
| Severe | 26 | 29 | 89.7 | 342 | 456 | 75.0 | 609 | 1,046 | 58.2 | 9,441 | 11,335 | 83.3 | 2,738 | 3,908 | 70.1 | 13,156 | 16,774 | 78.4 |
| Mild to moderate | 136 | 343 | 39.7 | 690 | 2,728 | 25.3 | 656 | 4,127 | 15.9 | 7,753 | 18,984 | 40.8 | 4,526 | 13,827 | 32.7 | 13,761 | 40,009 | 34.4 |
| **ICU admission<sup>a</sup>** |  |  |  |  |  |  |  |  |  |  |  |  |  |  |  |  |  |  |
| No | 164 | 374 | 43.9 | 1,087 | 3,239 | 33.6 | 1,305 | 5,216 | 25.0 | 23,622 | 36,987 | 63.9 | 7,803 | 18,303 | 42.6 | 33,981 | 64,119 | 53.0 |
| Yes | 25 | 31 | 80.6 | 161 | 163 | 98.8 | 218 | 232 | 94.0 | 909 | 1,004 | 90.5 | 213 | 261 | 81.6 | 1,526 | 1,691 | 90.2 |
| **Blood culture** |  |  |  |  |  |  |  |  |  |  |  |  |  |  |  |  |  |  |
| No | 118 | 307 | 38.4 | 854 | 2,810 | 30.4 | 1,146 | 4,808 | 23.8 | 17,900 | 30,258 | 59.2 | 5,513 | 14,899 | 37.0 | 25,531 | 53,082 | 48.1 |
| Yes | 71 | 98 | 72.4 | 394 | 592 | 66.6 | 377 | 640 | 58.9 | 6,631 | 7,733 | 85.7 | 2,503 | 3,665 | 68.3 | 9,976 | 12,728 | 78.4 |
| **Sputum culture** |  |  |  |  |  |  |  |  |  |  |  |  |  |  |  |  |  |  |
|  | No. with Rx | No. Total | % | No. with Rx | No. Total | % | No. with Rx | No. Total | % | No. with Rx | No. Total | % | No. with Rx | No. Total | % | No. with Rx | No. Total | % |
| No | 164 | 374 | 43.9 | 1,103 | 3,203 | 34.4 | 1,340 | 5,197 | 25.8 | 22,447 | 35,745 | 62.8 | 7,052 | 17,352 | 40.6 | 32,106 | 61,871 | 51.9 |
| Yes | 25 | 31 | 80.6 | 145 | 199 | 72.9 | 183 | 251 | 72.9 | 2,084 | 2,246 | 92.8 | 964 | 1,212 | 79.5 | 3,401 | 3,939 | 86.3 |
| **Urine culture** |  |  |  |  |  |  |  |  |  |  |  |  |  |  |  |  |  |  |
| No | 161 | 374 | 43.0 | 1,051 | 3,164 | 33.2 | 1,319 | 5,194 | 25.4 | 19,131 | 31,942 | 59.9 | 5,694 | 15,536 | 36.7 | 27,356 | 56,210 | 48.7 |
| Yes | 28 | 31 | 90.3 | 197 | 238 | 82.8 | 204 | 254 | 80.3 | 5,400 | 6,049 | 89.3 | 2,322 | 3,028 | 76.7 | 8,151 | 9,600 | 84.9 |
| **COVID-19 vaccine doses, number<sup>b</sup>** |  |  |  |  |  |  |  |  |  |  |  |  |  |  |  |  |  |  |
| 0 | 189 | 405 | 46.7 | 1,248 | 3,402 | 36.7 | 1,523 | 5,448 | 28.0 | 14,274 | 20,418 | 69.9 | 1,233 | 4,031 | 30.6 | 18,467 | 33,704 | 54.8 |
| 1 | NA | NA | NA | NA | NA | NA | 0 | 0 | 0.0 | 4,864 | 6,710 | 72.5 | 228 | 568 | 40.1 | 5,092 | 7,278 | 70.0 |
| 2 | NA | NA | NA | NA | NA | NA | 0 | 0 | 0.0 | 4,681 | 9,236 | 50.7 | 2,498 | 5,100 | 49.0 | 7,179 | 14,336 | 50.1 |
| 3 | NA | NA | NA | NA | NA | NA | 0 | 0 | 0.0 | 710 | 1,623 | 43.7 | 3,714 | 8,000 | 46.4 | 4,424 | 9,623 | 46.0 |
| 4 | NA | NA | NA | NA | NA | NA | 0 | 0 | 0.0 | 2 | 4 | 50.0 | 343 | 865 | 39.7 | 345 | 869 | 39.7 |
| **Antibacterial drugs** |  |  |  |  |  |  |  |  |  |  |  |  |  |  |  |  |  |  |
| No | 186 | 401 | 46.4 | 1,181 | 3,299 | 35.8 | 1,456 | 5,321 | 27.4 | 18,699 | 30,821 | 60.7 | 6,795 | 16,725 | 40.6 | 28,317 | 56,567 | 50.1 |
| Yes | 3 | 4 | 75.0 | 67 | 103 | 65.0 | 67 | 127 | 52.8 | 5,832 | 7,170 | 81.3 | 1,221 | 1,839 | 66.4 | 7,190 | 9,243 | 77.8 |
| **Respiratory drugs** |  |  |  |  |  |  |  |  |  |  |  |  |  |  |  |  |  |  |
|  | No.<br>with<br>Rx | No.<br>Total | % | No.<br>with<br>Rx | No.<br>Total | % | No.<br>with<br>Rx | No.<br>Total | % | No.<br>with<br>Rx | No.<br>Total | % | No.<br>with<br>Rx | No.<br>Total | % | No.<br>with<br>Rx | No.<br>Total | % |
| No | 189 | 404 | 46.8 | 1,217 | 3,358 | 36.2 | 1,501 | 5,395 | 27.8 | 21,040 | 33,968 | 61.9 | 7,312 | 17,526 | 41.7 | 31,259 | 60,651 | 51.5 |
| Yes | 0 | 1 | 0.0 | 31 | 44 | 70.5 | 22 | 53 | 41.5 | 3,491 | 4,023 | 86.8 | 704 | 1,038 | 67.8 | 4,248 | 5,159 | 82.3 |
| **Corticosteroids** |  |  |  |  |  |  |  |  |  |  |  |  |  |  |  |  |  |  |
| No | 189 | 405 | 46.7 | 1,225 | 3,368 | 36.4 | 1,504 | 5,422 | 27.7 | 20,992 | 33,858 | 62.0 | 7,408 | 17,641 | 42.0 | 31,318 | 60,694 | 51.6 |
| Yes | 0 | 0 | 0.0 | 23 | 34 | 67.6 | 19 | 26 | 73.1 | 3,539 | 4,133 | 85.6 | 608 | 923 | 65.9 | 4,189 | 5,116 | 81.9 |
| **Coronavirus antivirals** |  |  |  |  |  |  |  |  |  |  |  |  |  |  |  |  |  |  |
| No | 189 | 405 | 46.7 | 1,248 | 3,402 | 36.7 | 1,523 | 5,448 | 28.0 | 23,052 | 36,126 | 63.8 | 7,273 | 16,951 | 42.9 | 33,285 | 62,332 | 53.4 |
| Yes | 0 | 0 | 0.0 | 0 | 0 | 0.0 | 0 | 0 | 0.0 | 1,479 | 1,865 | 79.3 | 743 | 1,613 | 46.1 | 2,222 | 3,478 | 63.9 |
| **Hypertension** |  |  |  |  |  |  |  |  |  |  |  |  |  |  |  |  |  |  |
| No | 156 | 364 | 42.9 | 800 | 2,690 | 29.7 | 1,054 | 4,570 | 23.1 | 8,512 | 16,815 | 50.6 | 3,484 | 10,308 | 33.8 | 14,006 | 34,747 | 40.3 |
| Yes | 33 | 41 | 80.5 | 448 | 712 | 62.9 | 469 | 878 | 53.4 | 16,019 | 21,176 | 75.6 | 4,532 | 8,256 | 54.9 | 21,501 | 31,063 | 69.2 |
| **Diabetes mellitus** |  |  |  |  |  |  |  |  |  |  |  |  |  |  |  |  |  |  |
| No | 173 | 382 | 45.3 | 991 | 3,012 | 32.9 | 1,273 | 4,962 | 25.7 | 16,559 | 27,379 | 60.5 | 5,780 | 14,516 | 39.8 | 24,776 | 50,251 | 49.3 |
| Yes | 16 | 23 | 69.6 | 257 | 390 | 65.9 | 250 | 486 | 51.4 | 7,972 | 10,612 | 75.1 | 2,236 | 4,048 | 55.2 | 10,731 | 15,559 | 69.0 |
|  | No. with Rx | No. Total | % | No. with Rx | No. Total | % | No. with Rx | No. Total | % | No. with Rx | No. Total | % | No. with Rx | No. Total | % | No. with Rx | No. Total | % |
| **Cardiovascular disease** |  |  |  |  |  |  |  |  |  |  |  |  |  |  |  |  |  |  |
| No | 184 | 399 | 46.1 | 1,114 | 3,222 | 34.6 | 1,416 | 5,275 | 26.8 | 17,251 | 28,822 | 59.9 | 6,191 | 15,506 | 39.9 | 26,156 | 53,224 | 49.1 |
| Yes | 5 | 6 | 83.3 | 134 | 180 | 74.4 | 107 | 173 | 61.8 | 7,280 | 9,169 | 79.4 | 1,825 | 3,058 | 59.7 | 9,351 | 12,586 | 74.3 |
| **Malignancy** |  |  |  |  |  |  |  |  |  |  |  |  |  |  |  |  |  |  |
| No | 186 | 401 | 46.4 | 1,220 | 3,357 | 36.3 | 1,501 | 5,407 | 27.8 | 22,730 | 35,504 | 64.0 | 7,449 | 17,614 | 42.3 | 33,086 | 62,283 | 53.1 |
| Yes | 3 | 4 | 75.0 | 28 | 45 | 62.2 | 22 | 41 | 53.7 | 1,801 | 2,487 | 72.4 | 567 | 950 | 59.7 | 2,421 | 3,527 | 68.6 |
| **Chronic kidney disease** |  |  |  |  |  |  |  |  |  |  |  |  |  |  |  |  |  |  |
| No | 189 | 405 | 46.7 | 1,229 | 3,381 | 36.4 | 1,511 | 5,432 | 27.8 | 22,710 | 35,210 | 64.5 | 7,661 | 17,959 | 42.7 | 33,300 | 62,387 | 53.4 |
| Yes | 0 | 0 | 0.0 | 19 | 21 | 90.5 | 12 | 16 | 75.0 | 1,821 | 2,781 | 65.5 | 355 | 605 | 58.7 | 2,207 | 3,423 | 64.5 |
| **Chronic obstructive pulmonary disease** |  |  |  |  |  |  |  |  |  |  |  |  |  |  |  |  |  |  |
| No | 189 | 405 | 46.7 | 1,221 | 3,373 | 36.2 | 1,507 | 5,427 | 27.8 | 22,624 | 35,879 | 63.1 | 7,603 | 18,008 | 42.2 | 33,144 | 63,092 | 52.5 |
| Yes | 0 | 0 | 0.0 | 27 | 29 | 93.1 | 16 | 21 | 76.2 | 1,907 | 2,112 | 90.3 | 413 | 556 | 74.3 | 2,363 | 2,718 | 86.9 |
| **Chronic liver disease** |  |  |  |  |  |  |  |  |  |  |  |  |  |  |  |  |  |  |
| No | 188 | 403 | 46.7 | 1,233 | 3,367 | 36.6 | 1,500 | 5,398 | 27.8 | 23,624 | 36,806 | 64.2 | 7,788 | 18,175 | 42.9 | 34,333 | 64,149 | 53.5 |
|  | No. with Rx | No. Total | % | No. with Rx | No. Total | % | No. with Rx | No. Total | % | No. with Rx | No. Total | % | No. with Rx | No. Total | % | No. with Rx | No. Total | % |
| Yes | 1 | 2 | 50.0 | 15 | 35 | 42.9 | 23 | 50 | 46.0 | 907 | 1,185 | 76.5 | 228 | 389 | 58.6 | 1,174 | 1,661 | 70.7 |
| **Asthma** |  |  |  |  |  |  |  |  |  |  |  |  |  |  |  |  |  |  |
| No | 189 | 405 | 46.7 | 1,237 | 3,381 | 36.6 | 1,504 | 5,415 | 27.8 | 24,078 | 37,417 | 64.4 | 7,865 | 18,297 | 43.0 | 34,873 | 64,915 | 53.7 |
| Yes | 0 | 0 | 0.0 | 11 | 21 | 52.4 | 19 | 33 | 57.6 | 453 | 574 | 78.9 | 151 | 267 | 56.6 | 634 | 895 | 70.8 |
| **Rheumatic disease** |  |  |  |  |  |  |  |  |  |  |  |  |  |  |  |  |  |  |
| No | 189 | 405 | 46.7 | 1,247 | 3,400 | 36.7 | 1,520 | 5,441 | 27.9 | 24,355 | 37,739 | 64.5 | 7,950 | 18,448 | 43.1 | 35,261 | 65,433 | 53.9 |
| Yes | 0 | 0 | 0.0 | 1 | 2 | 50.0 | 3 | 7 | 42.9 | 176 | 252 | 69.8 | 66 | 116 | 56.9 | 246 | 377 | 65.3 |
| **Neutropenia** |  |  |  |  |  |  |  |  |  |  |  |  |  |  |  |  |  |  |
| No | 189 | 405 | 46.7 | 1,246 | 3,399 | 36.7 | 1,522 | 5,447 | 27.9 | 24,419 | 37,834 | 64.5 | 7,980 | 18,497 | 43.1 | 35,356 | 65,582 | 53.9 |
| Yes | 0 | 0 | 0.0 | 2 | 3 | 66.7 | 1 | 1 | 100.0 | 112 | 157 | 71.3 | 36 | 67 | 53.7 | 151 | 228 | 66.2 |
| **Organ transplant** |  |  |  |  |  |  |  |  |  |  |  |  |  |  |  |  |  |  |
| No | 189 | 405 | 46.7 | 1,247 | 3,399 | 36.7 | 1,522 | 5,447 | 27.9 | 24,436 | 37,834 | 64.6 | 7,978 | 18,506 | 43.1 | 35,372 | 65,591 | 53.9 |
| Yes | 0 | 0 | 0.0 | 1 | 3 | 33.3 | 1 | 1 | 100.0 | 95 | 157 | 60.5 | 38 | 58 | 65.5 | 135 | 219 | 61.6 |

<sup>a</sup>Inpatient admission characteristic.
<sup>b</sup>COVID-19 vaccines became available locally beginning in February 2021 (i.e., wave 4).

| Wave | 1-2 |  |  | 3 |  |  | 4 |  |  | 5E |  |  | 5L |  |  | Overall |  |  |
| --- | --- | --- | --- | --- | --- | --- | --- | --- | --- | --- | --- | --- | --- | --- | --- | --- | --- | --- |
|  | No.<br>with<br>Rx | No.<br>Total | % | No.<br>with<br>Rx | No.<br>Total | % | No.<br>with<br>Rx | No.<br>Total | % | No.<br>with<br>Rx | No.<br>Total | % | No.<br>with<br>Rx | No.<br>Total | % | No.<br>with<br>Rx | No.<br>Total | % |

Abbreviations: NA, not applicable; Rx, inpatient antibacterial drug prescription.

**Supplementary Table 4.** Antibacterial drug prescriptions according to British National Formulary (BNF) section for COVID-19 patients infected in the community in Hong Kong over five epidemic waves from 21 January 2020 through to 30 September 2022

| BNF Section | Description | DOT (days) | DOT (%) | DOT/1000 patients | DOT/1000 patient days |
| --- | --- | --- | --- | --- | --- |
| 5.1 | Antibacterial drugs | 416,031 | 100.0 | 6,321.7 | 550.5 |
| 5.1.1 | Penicillins | 234,893 | 56.5 | 3,569.3 | 310.8 |
| 5.1.1.1 | Benzylpenicillin and phenoxymethylpenicillin | 535 | 0.1 | 8.1 | 0.7 |
| 5.1.1.2 | Penicillinase-resistant penicillins | 1,390 | 0.3 | 21.1 | 1.8 |
| 5.1.1.3 | Broad-spectrum penicillins | 164,547 | 39.6 | 2,500.3 | 217.7 |
| 5.1.1.4 | Antipseudomonal penicillins | 68,421 | 16.4 | 1,039.7 | 90.5 |
| 5.1.2 | Cephalosporins, carbapenems, and other beta-lactams | 102,768 | 24.7 | 1,561.6 | 136.0 |
| 5.1.2.1 | Cephalosporins | 72,945 | 17.5 | 1,108.4 | 96.5 |
| 5.1.2.2 | Carbapenems | 29,802 | 7.2 | 452.8 | 39.4 |
| 5.1.2.3 | Other beta-lactam antibiotics | 21 | 0.0 | 0.3 | 0.0 |
| 5.1.3 | Tetracyclines | 13,606 | 3.3 | 206.7 | 18.0 |
| 5.1.4 | Aminoglycosides | 2,304 | 0.6 | 35.0 | 3.0 |
| 5.1.5 | Macrolides | 3,694 | 0.9 | 56.1 | 4.9 |
| 5.1.6 | Clindamycin and lincomycin | 545 | 0.1 | 8.3 | 0.7 |
| 5.1.7 | Some other antibacterials | 15,107 | 3.6 | 229.6 | 20.0 |
| 5.1.8 | Sulfonamides and trimethoprim | 5,453 | 1.3 | 82.9 | 7.2 |
| 5.1.9 | Antituberculosis drugs | 10,025 | 2.4 | 152.3 | 13.3 |
| 5.1.10 | Antileprotic drugs | 429 | 0.1 | 6.5 | 0.6 |
| 5.1.11 | Metronidazole and tinidazole | 4,747 | 1.1 | 72.1 | 6.3 |
| 5.1.12 | Quinolones | 21,275 | 5.1 | 323.3 | 28.2 |
| 5.1.13 | Urinary-tract infections | 1,185 | 0.3 | 18.0 | 1.6 |

| **BNF**<br>**Section** | **Description** | **DOT**<br>**(days)** | **DOT**<br>**(%)** | **DOT/1000**<br>**patients** | **DOT/1000 patient**<br>**days** |
| --- | --- | --- | --- | --- | --- |

Abbreviations: BNF, British National Formulary; DOT, days of therapy.

**Supplementary Figure 3.**
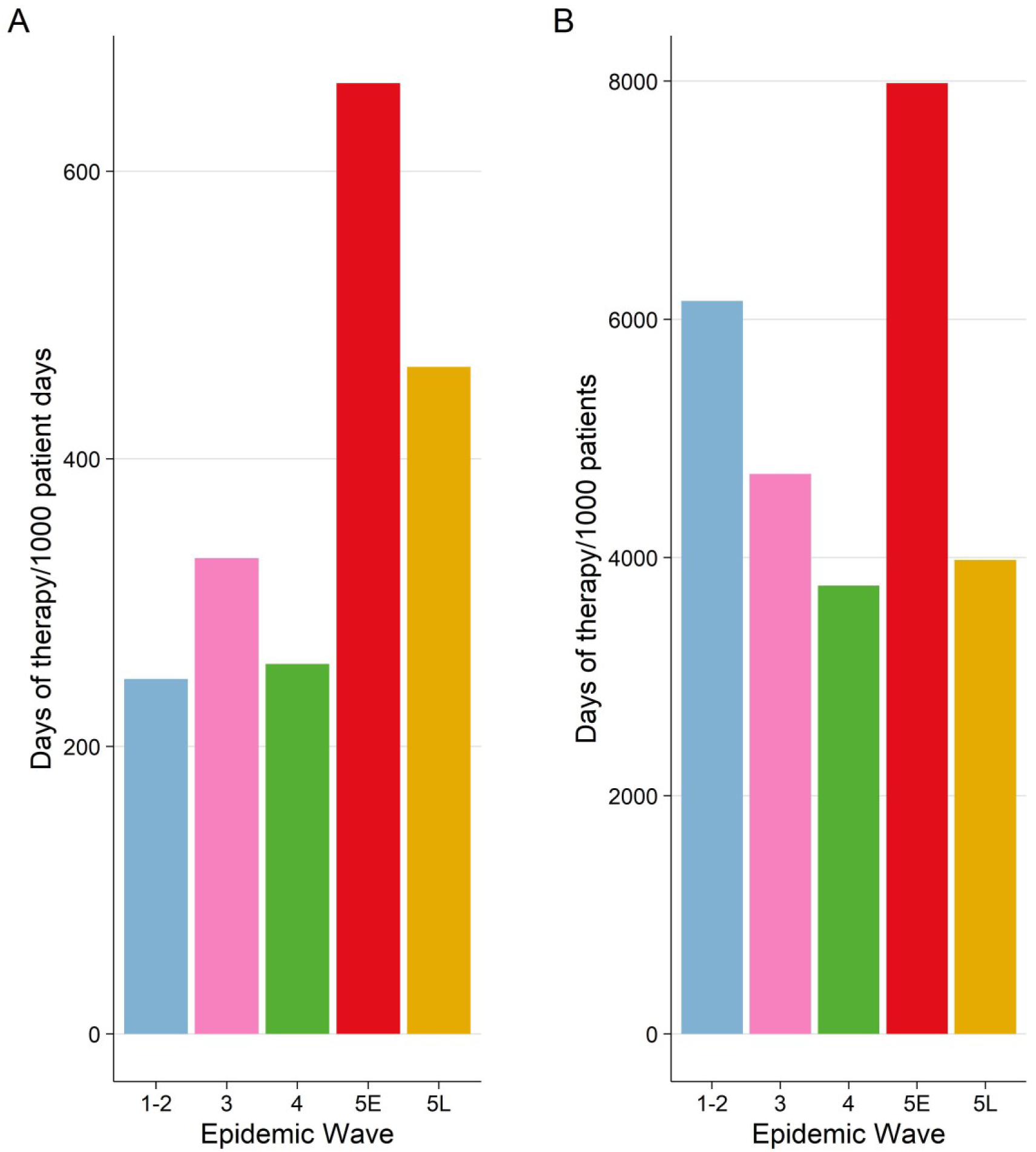
Antibacterial drug use by wave as measured by days of therapy/1000 patient days (**A**) and days of therapy/1000 patients (**B**)

**Supplementary Table 5.** Days of antibacterial therapy, initiation day of antibacterial therapy, and rates of use for each antibacterial drug in COVID-19 patients included in the analysis over 5 epidemic waves in Hong Kong

| Rank | BNF | Antibacterial drug | DOT (days) | DOT (%) | DOT, median [IQR] | Initiation day, median [IQR] | Days of therapy/1000 patients | Days of therapy/1000 patient days |
| --- | --- | --- | --- | --- | --- | --- | --- | --- |
| 1 | 5.1.1.3 | Amoxicillin/clavulanic acid | 161,694 | 38.9 | 6.0 [3.0, 8.0] | 0.0 [0.0, 1.0] | 2,456.98 | 213.95 |
| 2 | 5.1.1.4 | Piperacillin/tazobactam | 67,915 | 16.3 | 7.0 [4.0, 9.0] | 3.0 [0.0, 7.0] | 1,031.99 | 89.86 |
| 3 | 5.1.2.1 | Ceftriaxone | 52,936 | 12.7 | 5.0 [3.0, 8.0] | 0.0 [0.0, 1.0] | 804.38 | 70.04 |
| 4 | 5.1.2.2 | Meropenem | 26,962 | 6.5 | 8.0 [4.0, 11.0] | 8.0 [3.0, 14.0] | 409.69 | 35.68 |
| 5 | 5.1.12 | Levofloxacin | 19,042 | 4.6 | 6.0 [3.0, 8.0] | 3.0 [0.0, 11.0] | 289.35 | 25.20 |
| 6 | 5.1.7 | Vancomycin | 12,738 | 3.1 | 7.0 [3.0, 11.0] | 11.0 [4.0, 21.0] | 193.56 | 16.85 |
| 7 | 5.1.3 | Doxycycline | 11,871 | 2.9 | 6.0 [4.0, 8.0] | 1.0 [0.0, 3.0] | 180.38 | 15.71 |
| 8 | 5.1.2.1 | Cefoperazone/sulbactam | 7,350 | 1.8 | 7.0 [4.0, 9.0] | 8.0 [3.0, 16.0] | 111.69 | 9.73 |
| 9 | 5.1.8 | Trimethoprim/sulfamethoxazole | 5,371 | 1.3 | 7.0 [4.0, 13.0] | 1.0 [0.0, 13.0] | 81.61 | 7.11 |
| 10 | 5.1.11 | Metronidazole | 4,747 | 1.1 | 5.0 [3.0, 8.0] | 2.0 [0.0, 9.8] | 72.13 | 6.28 |
| 11 | 5.1.2.1 | Ceftazidime | 3,230 | 0.8 | 7.0 [4.0, 10.0] | 10.0 [3.0, 20.0] | 49.08 | 4.27 |
| 12 | 5.1.9 | Isoniazid | 3,090 | 0.7 | 10.0 [5.5, 19.0] | 1.0 [0.0, 5.0] | 46.95 | 4.09 |
| 13 | 5.1.9 | Rifampicin | 3,081 | 0.7 | 9.0 [5.0, 19.0] | 2.0 [0.0, 7.0] | 46.82 | 4.08 |
| 14 | 5.1.2.1 | Cefuroxime | 2,848 | 0.7 | 5.0 [3.0, 8.0] | 4.0 [1.0, 10.0] | 43.28 | 3.77 |
| 15 | 5.1.5 | Azithromycin | 2,728 | 0.7 | 3.0 [1.0, 4.0] | 0.0 [0.0, 1.0] | 41.45 | 3.61 |
| 16 | 5.1.2.1 | Cefepime | 2,591 | 0.6 | 7.0 [4.0, 10.0] | 10.0 [5.0, 19.0] | 39.37 | 3.43 |
| 17 | 5.1.2.2 | Ertapenem | 2,383 | 0.6 | 7.0 [4.0, 8.0] | 6.0 [4.0, 17.0] | 36.21 | 3.15 |
| 18 | 5.1.2.1 | Cefazolin | 2,088 | 0.5 | 2.0 [1.0, 3.0] | 0.0 [0.0, 3.0] | 31.73 | 2.76 |
| 19 | 5.1.9 | Ethambutol | 2,088 | 0.5 | 11.0 [6.0, 21.0] | 2.0 [1.0, 8.0] | 31.73 | 2.76 |
| 20 | 5.1.12 | Ciprofloxacin | 1,938 | 0.5 | 4.0 [3.0, 7.0] | 3.0 [0.0, 9.0] | 29.45 | 2.56 |
| 21 | 5.1.1.3 | Ampicillin | 1,844 | 0.4 | 4.0 [2.0, 5.0] | 0.0 [0.0, 1.0] | 28.02 | 2.44 |
| 22 | 5.1.9 | Pyrazinamide | 1,597 | 0.4 | 11.0 [6.0, 19.0] | 2.0 [1.0, 12.0] | 24.27 | 2.11 |
| 23 | 5.1.3 | Minocycline | 1,537 | 0.4 | 5.0 [3.0, 8.0] | 2.0 [0.0, 19.0] | 23.36 | 2.03 |
| 24 | 5.1.1.2 | Cloxacillin | 1,390 | 0.3 | 5.0 [3.0, 8.0] | 5.0 [1.0, 12.0] | 21.12 | 1.84 |
| 25 | 5.1.2.1 | Cefotaxime | 1,194 | 0.3 | 5.0 [3.0, 6.0] | 0.0 [0.0, 1.0] | 18.14 | 1.58 |
| 26 | 5.1.13 | Nitrofurantoin | 1,185 | 0.3 | 5.0 [3.0, 7.0] | 7.0 [4.0, 17.5] | 18.01 | 1.57 |
| 27 | 5.1.4 | Gentamicin | 1,137 | 0.3 | 3.0 [2.0, 6.0] | 2.0 [0.0, 14.0] | 17.28 | 1.50 |
| 28 | 5.1.4 | Amikacin | 1,117 | 0.3 | 2.0 [1.0, 6.0] | 6.5 [1.0, 18.0] | 16.97 | 1.48 |
| 29 | 5.1.7 | Linezolid | 825 | 0.2 | 7.0 [4.0, 9.0] | 13.0 [6.2, 22.0] | 12.54 | 1.09 |
| 30 | 5.1.5 | Clarithromycin | 738 | 0.2 | 4.0 [2.0, 7.0] | 1.0 [0.0, 6.0] | 11.21 | 0.98 |
| 31 | 5.1.7 | Rifaximin | 719 | 0.2 | 9.0 [5.0, 15.5] | 0.0 [0.0, 2.0] | 10.93 | 0.95 |
| 32 | 5.1.6 | Clindamycin | 545 | 0.1 | 6.0 [3.0, 9.0] | 5.0 [0.0, 9.8] | 8.28 | 0.72 |
| 33 | 5.1.7 | Colistin | 509 | 0.1 | 8.0 [5.0, 16.0] | 21.5 [12.2, 29.8] | 7.73 | 0.67 |
| 34 | 5.1.1.3 | Amoxicillin | 494 | 0.1 | 4.0 [2.0, 7.0] | 3.0 [0.0, 8.0] | 7.51 | 0.65 |
| 35 | 5.1.1.3 | Ampicillin/sulbactam | 459 | 0.1 | 7.0 [4.0, 13.0] | 13.0 [7.0, 32.0] | 6.97 | 0.61 |
| 36 | 5.1.2.2 | Imipenem | 457 | 0.1 | 6.0 [3.0, 9.0] | 3.0 [1.0, 6.0] | 6.94 | 0.60 |
| 37 | 5.1.1.4 | Ticarcillin/clavulanic acid | 383 | 0.1 | 7.0 [3.0, 11.0] | 17.0 [11.5, 34.5] | 5.82 | 0.51 |
| 38 | 5.1.10 | Clofazimine | 376 | 0.1 | 2.0 [1.0, 3.0] | 1.0 [0.0, 2.0] | 5.71 | 0.50 |
| 39 | 5.1.2.1 | Ceftazidime/avibactam | 364 | 0.1 | 8.0 [4.0, 11.5] | 13.0 [7.0, 20.0] | 5.53 | 0.48 |
| 40 | 5.1.1.1 | Benzylpenicillin | 353 | 0.1 | 2.0 [2.0, 6.0] | 0.0 [0.0, 2.0] | 5.36 | 0.47 |
| 41 | 5.1.12 | Moxifloxacin | 295 | 0.1 | 7.0 [3.8, 12.5] | 8.0 [1.0, 22.0] | 4.48 | 0.39 |
| 42 | 5.1.5 | Erythromycin | 228 | 0.1 | 7.0 [4.0, 12.0] | 1.0 [0.0, 5.0] | 3.46 | 0.30 |
| 43 | 5.1.7 | Daptomycin | 208 | 0.0 | 7.0 [2.5, 15.0] | 22.0 [13.5, 27.5] | 3.16 | 0.28 |
| 44 | 5.1.3 | Tigecycline | 179 | 0.0 | 10.0 [5.0, 12.0] | 17.0 [8.5, 29.5] | 2.72 | 0.24 |
| 45 | 5.1.1.1 | Phenoxymethylpenicillin | 161 | 0.0 | 4.5 [2.8, 7.0] | 0.5 [0.0, 2.0] | 2.45 | 0.21 |
| 46 | 5.1.2.1 | Ceftaroline | 139 | 0.0 | 9.0 [7.0, 10.2] | 18.5 [10.8, 25.0] | 2.11 | 0.18 |
| 47 | 5.1.9 | Rifabutin | 134 | 0.0 | 17.0 [12.0, 17.0] | 1.0 [1.0, 2.0] | 2.04 | 0.18 |
| 48 | 5.1.1.4 | Piperacillin | 123 | 0.0 | 8.0 [6.0, 9.5] | 11.0 [5.5, 22.0] | 1.87 | 0.16 |
| 49 | 5.1.7 | Fusidic acid | 85 | 0.0 | 9.0 [5.0, 19.0] | 24.0 [21.0, 37.0] | 1.29 | 0.11 |
| 50 | 5.1.8 | Trimethoprim | 82 | 0.0 | 3.0 [1.0, 4.0] | 1.0 [0.0, 4.0] | 1.25 | 0.11 |
| 51 | 5.1.2.1 | Cephalexin | 62 | 0.0 | 3.5 [1.8, 6.0] | 7.5 [1.0, 11.2] | 0.94 | 0.08 |
| 52 | 5.1.1.3 | Sultamicillin | 56 | 0.0 | 4.0 [2.2, 5.8] | 7.0 [2.5, 13.0] | 0.85 | 0.07 |
| 53 | 5.1.10 | Dapsone | 53 | 0.0 | 4.5 [4.0, 8.8] | 0.5 [0.0, 1.5] | 0.81 | 0.07 |
| 54 | 5.1.2.1 | Cefpodoxime | 51 | 0.0 | 1.5 [1.0, 3.0] | 4.0 [1.0, 6.0] | 0.77 | 0.07 |
| 55 | 5.1.2.1 | Cefoxitin | 38 | 0.0 | 17.0 [9.5, 18.0] | 1.0 [0.5, 20.0] | 0.58 | 0.05 |
| 56 | 5.1.2.1 | Ceftolozane/tazobactam | 38 | 0.0 | 13.0 [8.5, 17.0] | 36.0 [23.0, 39.0] | 0.58 | 0.05 |
| 57 | 5.1.4 | Tobramycin | 37 | 0.0 | 18.5 [13.8, 23.2] | 1.5 [0.8, 2.2] | 0.56 | 0.05 |
| 58 | 5.1.9 | Streptomycin | 35 | 0.0 | 4.0 [4.0, 8.0] | 4.0 [1.0, 10.0] | 0.53 | 0.05 |
| 59 | 5.1.7 | Fosfomycin | 23 | 0.0 | 1.0 [1.0, 2.8] | 9.0 [4.2, 14.8] | 0.35 | 0.03 |
| 60 | 5.1.2.3 | Aztreonam | 21 | 0.0 | 9.0 [5.0, 10.0] | 2.0 [1.0, 8.5] | 0.32 | 0.03 |
| 61 | 5.1.1.1 | Benzathine benzylpenicillin | 21 | 0.0 | 10.5 [9.2, 11.8] | 18.0 [9.0, 27.0] | 0.32 | 0.03 |
| 62 | 5.1.3 | Tetracycline | 19 | 0.0 | 4.0 [3.0, 4.0] | 6.0 [2.0, 26.0] | 0.29 | 0.03 |
| 63 | 5.1.4 | Neomycin | 13 | 0.0 | 1.0 [1.0, 1.0] | 0.0 [0.0, 0.0] | 0.20 | 0.02 |
| 64 | 5.1.2.1 | Cefaclor | 11 | 0.0 | 11.0 [11.0, 11.0] | 5.0 [5.0, 5.0] | 0.17 | 0.01 |
| 65 | 5.1.2.1 | Cefiderocol | 5 | 0.0 | 2.5 [2.2, 2.8] | 11.0 [9.0, 13.0] | 0.08 | 0.01 |

**Supplementary Figure 4.**
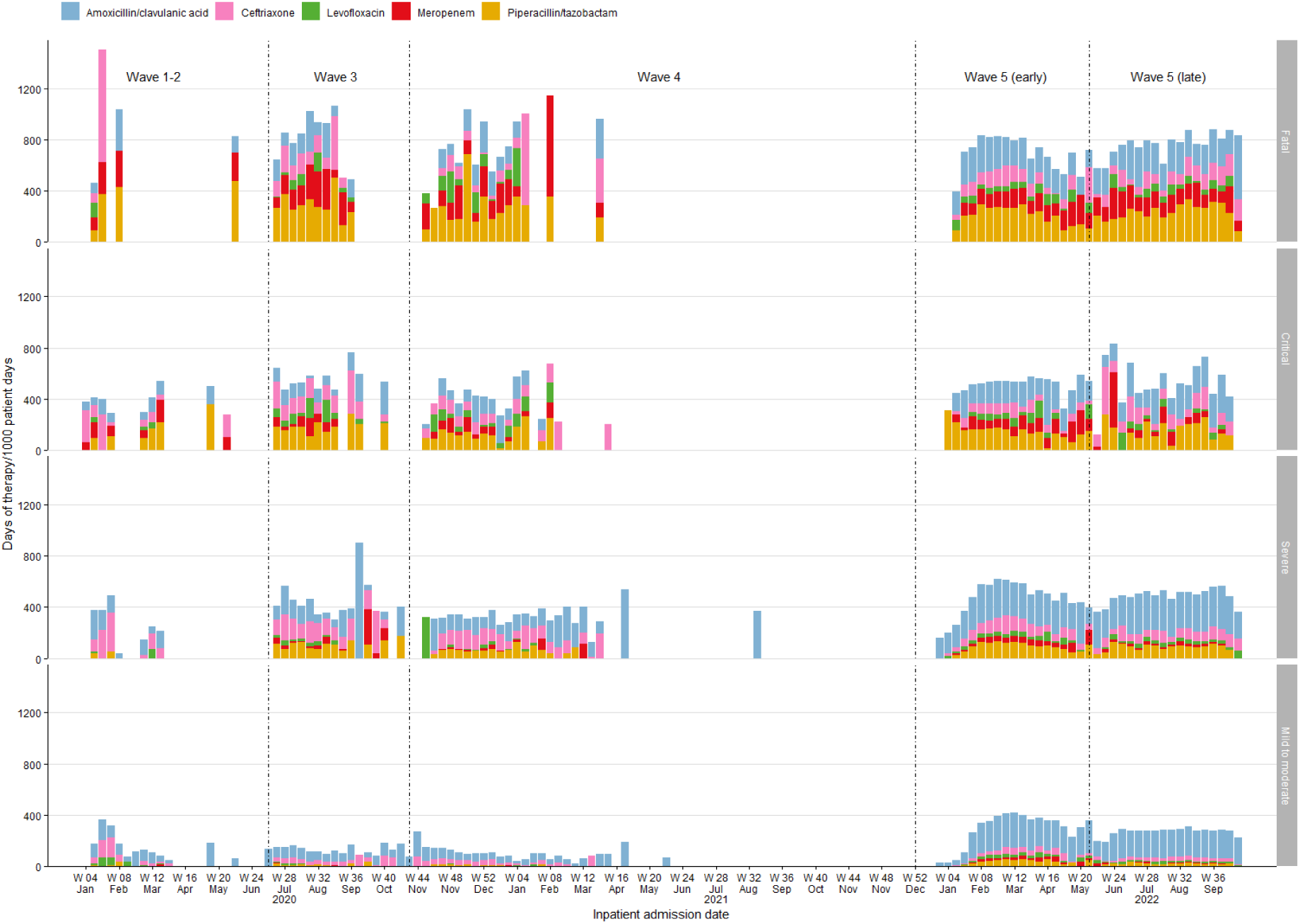
Weekly rate of use for the 5 most prescribed antibacterial drugs (days of therapy/1000 patient days) by COVID-19 disease severity

**Supplementary Figure 5.**
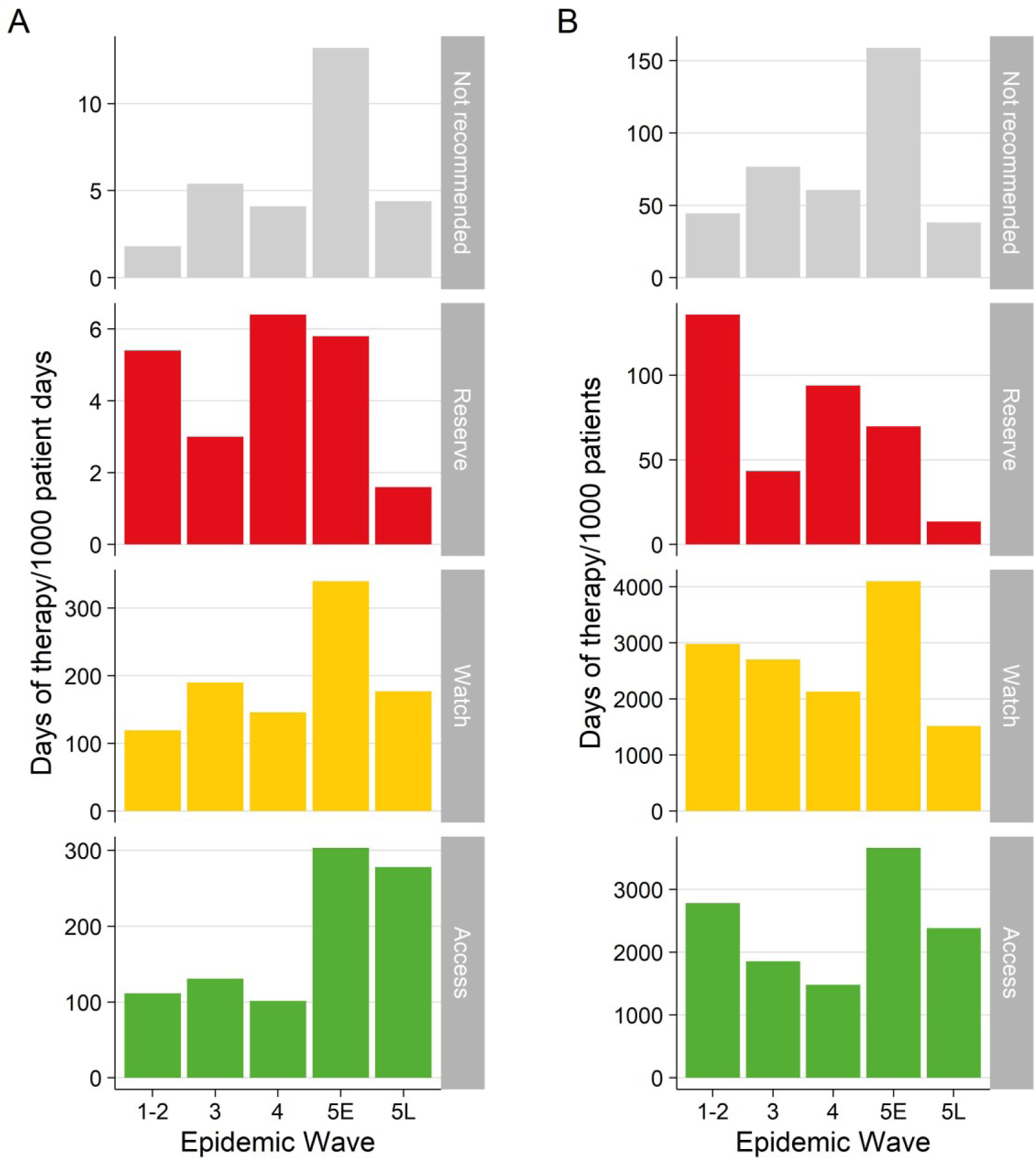
Use of antibacterial drugs according to the WHO AWaRe classification and by epidemic wave measured by days of therapy/1000 patient days (**A**) and days of therapy/1000 patients (**B**)

**Supplementary Figure 6.**
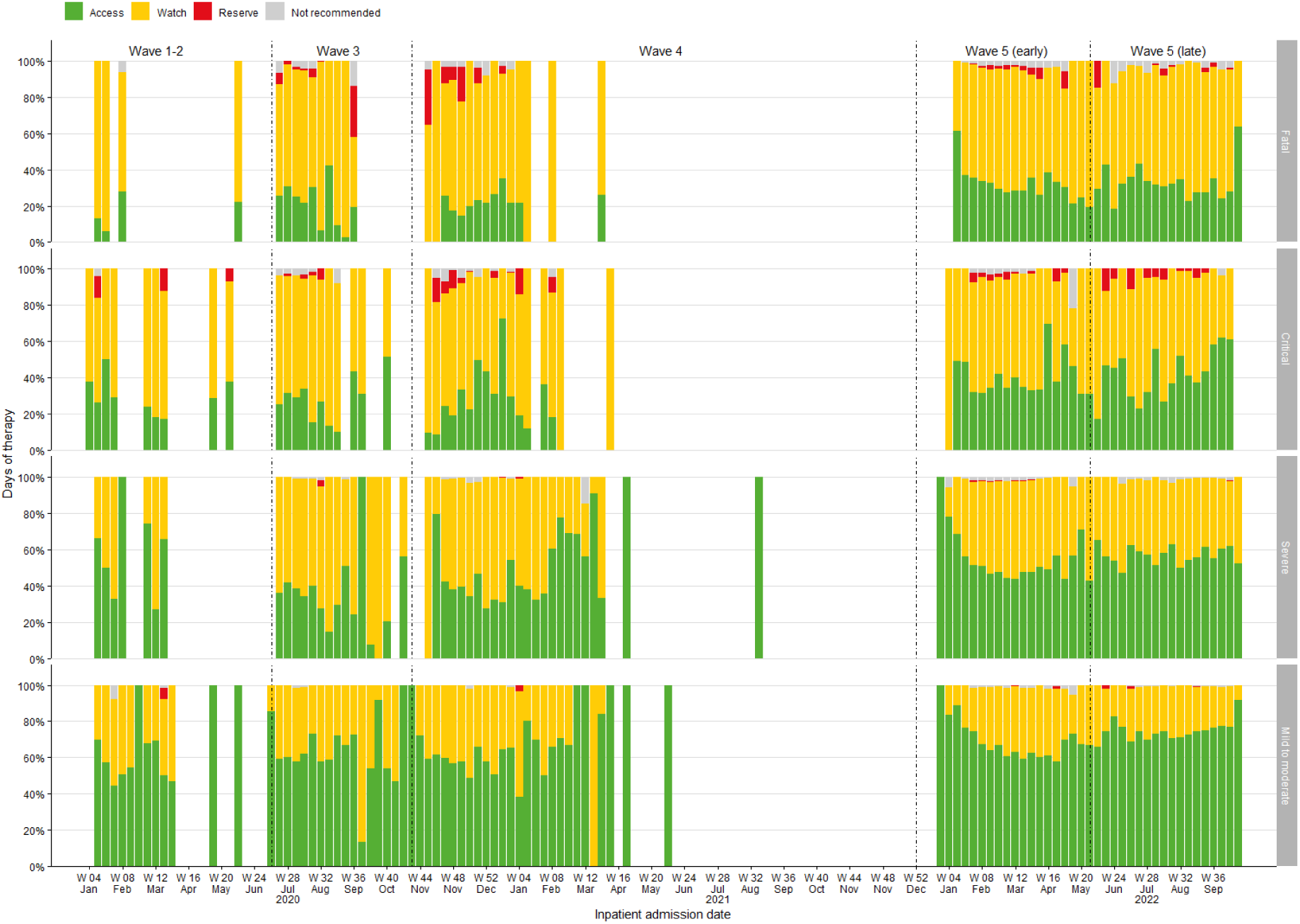
Weekly proportion of days of therapy by WHO AWaRe group and COVID-19 disease severity

